# Prevalence, duration, and clinical implications of Continuous Glucose Monitor (CGM) measurement limit capping in type 1 diabetes

**DOI:** 10.64898/2026.05.13.26353094

**Authors:** John F Mulley

**Affiliations:** School of Environmental and Natural Sciences, Bangor University, Bangor, Gwynedd, LL57 2UW, UK

**Keywords:** Continuous glucose monitoring, Type 1 diabetes, Sensor measurement limits, Time in range, Hyperglycaemia

## Abstract

**Aims:** CGM devices report glucose only within fixed limits (typically 40-400 mg/dL; 2.2-22.2 mmol/L), truncating extreme values to a boundary (‘capping’). We characterised prevalence, duration, and consequences of capping in type 1 diabetes trial data.

**Materials and Methods:** We analysed 46,990,617 CGM readings from 948 participants across four publicly available clinical trial datasets (Dexcom G4 Platinum or G6 sensors). Capping prevalence, run duration, and associations with age, HbA1c and sex were characterised across all datasets. In the 77 participants of the Replace-BG trial CGM-plus-blood glucose monitor (BGM) arm, CGM-derived metrics were compared with contemporaneous BGM measurements across 1,162 non-overlapping 14-day windows.

**Results:** Between 93.5% and 100% of participants had at least one capped reading, and capped values comprised 0.47–0.98% of all readings. In the three datasets for which duration could be calculated, over 70% of upper-cap runs exceeded 15 minutes and over one third exceeded 60 minutes. Upper-limit capping was inversely associated with age (Spearman ρ −0.20 to −0.47, p≤0.002) in three of the datasets, and positively associated with baseline HbA1c (ρ 0.39–0.62, p<0.001) in all four datasets. A within-participant analysis showed that capping burden did not predict CGM-BGM divergence in any summary metric (all p>0.2), and a systematic CGM-BGM offset in mean glucose and time in range (TIR) reflected the physiological lag between blood and interstitial fluid rather than capping artefact.

**Conclusions:** Sensor limit capping is near-universal in type 1 diabetes, produces sustained periods of right-censored glucose data disproportionately affecting younger patients, and does not substantially distort standard summary metrics at the population level. Clinicians and trialists should be aware that CGM data can confirm extreme glucose events but cannot quantify their severity.

**Novelty statement:** *What is already known about this subject?:* CGM devices operate within fixed measurement limits (40-400 mg/dL), and truncate extreme values. Glucose distributions in type 1 diabetes are right-skewed, with frequent hyperglycaemic excursions approaching upper sensor limits.

*What has this study found?:* Sensor limit capping is near-universal (93.5-100% of participants) and produces sustained periods (often exceeding 60 minutes) during which CGM cannot quantify true hyperglycaemia severity. Capping is inversely associated with age, positively associated with HbA1c, and does not substantially distort summary glucose metrics.

*What are the implications of this study?:* CGM records can confirm that extreme glucose events occurred but cannot quantify their severity. Clinicians, trialists, and guideline authors should account for this when interpreting extreme-range metrics, particularly in younger patients and paediatric trial populations.

## Introduction

Continuous glucose monitoring (CGM) has become central to the management of type 1 diabetes and the conduct of clinical trials in this population^1–3^. CGM devices are biosensors which use subcutaneous glucose oxidase-coated probes to measure glucose levels in the interstitial fluid between cells rather than in the blood directly, and CGM-derived metrics such as time in range (TIR), glucose management indicator (GMI), and time above or below range (TAR/TBR) now feature in National Institute for Health and Care Excellence (NICE) guidance, international consensus targets, and as primary endpoints in major trials^4,5^. Despite this, an in-built limitation of CGM devices has received little systematic attention: they can report glucose levels within interstitial fluid only within a fixed measurement window, typically 40-400 mg/dL (2.2-22.2 mmol/L), and so can only report that glucose has reached or exceed this limit, not the actual value. This introduces a form of statistical censoring in which the true severity of extreme glucose events is unknowable from sensor records alone.

Given the right-skewed glucose distributions typical of type 1 diabetes, where hyperglycaemic events are more frequent and more extreme than hypoglycaemic events^6^, upper sensor limits are particularly vulnerable to truncation. The consequences for clinical practice and research depends on how often capping occurs, how long it persists, and whether it distorts CGM-derived metrics on which conclusions are based. To our knowledge, these questions have not yet been systematically addressed, and the phenomenon of CGM measurement limit capping has not previously been characterised as distinct from sensor *accuracy*, which is based on mean absolute relative difference (MARD) between CGM and blood glucose reference values.

We therefore analysed four large, publicly available CGM datasets from type 1 diabetes trials to characterise capping prevalence and duration, its associations with patient age, sex, and HbA1c, and its impact on several commonly used glucose metrics.

## Materials and Methods

### Datasets

Four large-scale, publicly available clinical trial datasets were downloaded from the JAEB Center for Health Research (https://public.jaeb.org/datasets/diabetes): Aleppo et al. (2017^7^, *Replace-BG trial* NCT02258373 [version: Replace-BG Dataset]); Brown et al. (2019^8^, *The International Diabetes Closed Loop (iDCL) trial* NCT03563313 [version: DCLP3 Public Dataset - Release 3 - 2022-08-04]); Lynch et al. (2022^9^, *The Insulin-Only Bionic Pancreas Pivotal Trial* NCT04200313 [version: IOBP2 RCT Public Dataset]); and Wadwa et al. (2023^10^, *The Pediatric Artificial Pancreas (PEDAP) Trial* NCT04796779 [version: PEDAP Public Dataset - Release 5 - 2025-05-12]), hereafter “Aleppo”, “Brown”, “Lynch”, and “Wadwa”. These were selected as the largest individual-level CGM datasets from type 1 diabetes trials in the public domain, spanning paediatric to adult age ranges and using Dexcom sensors with the standard 40–400 mg/dL measurement range. The Brown, Lynch, and Wadwa studies used the Dexcom G6 CGM, and Aleppo used the Dexcom G4 Platinum. Each study had institutional ethics approval, and so the present pooled data analysis required no additional review. The analyses, content, and conclusions presented herein are solely the responsibility of the author and have not been reviewed or approved by authors or sponsors of the original trials.

### Data deduplication

Duplicate records were removed in two steps: exact duplicates (n=955) and timestamp conflicts (n=843,810), the latter largely arising from repeated Dexcom Clarity uploads in Wadwa. After deduplication, 46,990,617 records from 948 participants were available for analysis.

### Capping prevalence and spike ratios

To establish how capped reads are reported in these large-scale CGM datasets, we first sought to identify whether capped reads were reported at the device detection limit, or some arbitrary value adjacent to it. Readings ≤40 mg/dL (≤2.2 mmol/L) and ≥400 mg/dL (≥22.2 mmol/L) were quantified as proportions of all readings and of participants. A spike ratio was computed at each limit as the count of readings at the modal cap value (39 or 401 mg/dL) divided by the mean count of the two adjacent 5 mg/dL bins, and a spike ratio substantially greater than 1.0 is consistent with truncation at the sensor limit rather than physiological clustering. Beyond-limit readings present in Lynch and Aleppo were retained at their recorded values and not truncated to sensor limit values for analysis.

### Capping run analysis

Next, we quantified whether these capped readings were sporadic, one-off events, or part of longer runs of hypo- and hyperglycaemia. Consecutive capped readings within each participant’s data were identified as discrete runs at lower (≤40 mg/dL) and upper (≥400 mg/dL) limits. Run duration was calculated from first to last reading plus one 5-minute interval at thresholds of ≥15 minutes (≥3 readings), ≥30 minutes (≥6 readings), and ≥60 minutes (≥12 readings).

### Sensitivity analysis

To assess whether upper-limit capping metrics differed between the Dexcom G4 Platinum (Aleppo) and Dexcom G6 (Brown, Lynch, Wadwa) sensors, per-participant capping statistics were compared (G4 vs G6) using Mann-Whitney U tests with rank-biserial correlation as a measure of effect size. Internal consistency across the three G6 datasets was assessed using the Kruskal-Wallis test.

### Associations with age and sex

Per-participant capping metrics (percentage of readings at each limit, total time, longest run) were linked to demographic data from trial source files. Sex differences were assessed using Mann-Whitney U tests within each dataset; continuous age associations were assessed by Spearman rank correlation; and age group comparisons (<18, 18-39, 40-59, ≥60 years) by Kruskal-Wallis tests. Wadwa only enrolled participants ≤18 and was excluded from age group comparisons. These analyses are exploratory, and no correction for multiple comparisons was applied.

### Association between baseline HbA1c and upper-limit capping burden

To investigate whether participants with poorer baseline glycaemic control experienced greater sensor measurement limit capping, we computed Spearman rank correlations between baseline HbA1c and three per-participant upper-limit capping metrics: (i) percentage of CGM readings at the upper cap; (ii) total time spent at the upper cap; and (iii) the longest single upper-cap run. Baseline HbA1c was taken from the randomisation visit central laboratory measurement (Brown = *DiabLocalHbA1c_a*.*txt* file, Lynch = *tblALabHbA1c*.*csv*) or randomisation visit local laboratory measurement (Aleppo = *HLocalHbA1c*.*txt*). For Wadwa, the central laboratory HbA1c measured at randomisation (mean 7.6 ± 1.1%) was used (*STASampleResults*.*txt* file). Correlations were reported as Spearman ρ with 95% confidence intervals derived by Fisher z-transformation and two-sided p-values. All four datasets were analysed separately and results pooled across the combined cohort (n=945 participants with complete HbA1c and capping data).

### CGM versus blood glucose monitoring (BGM) comparison

Paired CGM and BGM data were analysed in the Aleppo CGM+BGM arm (n=77), based on contemporaneous reference measurements from the ContourOne BGM (measurement range 20-600 mg/dL, 1.1-33.3 mmol/L). Each BGM reading was matched to the nearest CGM reading within ±5 minutes. Lower-cap CGM readings (≤40 mg/dL) were considered confirmed by the paired BGM if the BGM was below 54 mg/dL (3.0 mmol/L), consistent with recognised thresholds for clinically significant hypoglycaemia^4,5,11^; upper-cap readings (≥400 mg/dL) were confirmed if BGM ≥400 mg/dL (22.2 mmol/L).

Data were divided into non-overlapping 14-day windows with a minimum 70% CGM data coverage and ≥1 BGM reading. Within each window, mean glucose, standard deviation (SD), coefficient of variation (CV), GMI (as 3.31 + 0.02392 x mean glucose mg/dL^12^), TIR 70-180 mg/dL, TAR >180 and >250 mg/dL, and TBR <70 and <54 mg/dL were calculated separately from CGM and BGM data. CGM-BGM differences (CGM minus BGM) were tested using the Wilcoxon signed-rank test.

The primary test for whether capping specifically caused metric divergence was a within-participant analysis in the 65 participants with sufficient windows to form above- and below-median capping halves. Comparing metric divergence between higher- and lower-cap windows within each participant controlled for the stable physiological interstitial-capillary glucose offset and for between-person differences in CGM-BGM agreement. Between-window tertile comparisons (pooling windows across participants) are reported as secondary analyses, with the caveat that pooled windows violate independence assumptions.

## Results

### Capping is near-universal and produces artefactual read accumulation

After deduplication, 46,990,617 CGM readings from 948 participants were available for analysis (Table 1, Supplementary Table S1). We find that between 93.5% and 100% of participants had at least one capped reading. Capped readings comprised 0.47-0.98% of all observations, with lower-limit readings (≤40 mg/dL; ≤2.2 mmol/L) present in 85.1-99.0% of participants, and upper-limit readings (≥400 mg/dL; ≥22.2 mmol/L) in 72.6-92.2%.

**Table 1.** Participant counts, baseline Hba1c, and CGM data characteristics for the four datasets after deduplication.

| Dataset | N | Baseline Hba1c (mean + SD, mmol/l and %) | Total readings | Min glucose reading (mg/dL) | Max glucose reading (mg/dL) | N readings ≤40 mg/dl [%] | N participants with readings ≤40 mg/dL [%] | N readings ≥400 mg/dl [%] | N participants with readings ≥400 mg/dL [%] | N out of bounds readings [%] | N participants with out of bounds readings [%] |
| --- | --- | --- | --- | --- | --- | --- | --- | --- | --- | --- | --- |
| Aleppo 2017<sup>7</sup> | 226 | 53 ± 8 (7.0 ± 0.7%) | 14948968 | 25 | 600 | 29024 [0.194] | 221 [97.8] | 49508 [0.331] | 201 [88.9] | 78532 [0.525] | 224 [99.1] |
| Brown 2019<sup>8</sup> | 168 | 57 ± 10 (7.4 ± 0.9%) | 7480842 | 39 | 401 | 4875 [0.065] | 143 [85.1] | 30608 [0.409] | 122 [72.6] | 35483 [0.474] | 157 [93.5] |
| Lynch 2022<sup>9</sup> | 451 | 61 ± 11 (7.7 ± 1.0%) | 18159404 | 12 | 654 | 76229 [0.42] | 401 [88.9] | 101160 [0.557] | 391 [86.7] | 177389 [0.977] | 443 [98.2] |
| Wadwa 2023<sup>10</sup> | 103 | 60 ± 12 (7.6 ± 1.1%) | 6401403 | 39 | 401 | 8360 [0.131] | 102 [99] | 32203 [0.503] | 95 [92.2] | 40563 [0.634] | 103 [100] |

In Brown and Wadwa, all glucose values were confined to the 39-401 mg/dL range, consistent with device-level data censoring at the measurement limits. Lynch and Aleppo contained small proportions of beyond-limit readings that were spread across participants and the duration of the study, which we could not attribute to a particular cause. However, in all four datasets, values at exactly 39 and 401 mg/dL were far more frequent than would be expected from the surrounding distribution (Figure 1, Supplementary Table S2a/b). Lower-limit spike ratios were 2.4-2.9x the mean of adjacent bins; upper-limit ratios were 7.2-21.8x, consistent with more pronounced truncation of extreme hyperglycaemia. These near-zero adjacent bin counts are contrary to what would be expected from a physiologically continuous distribution and confirm the artefactual accumulation of sensor readings at the device limits. Indeed, upper-limit capping was so pronounced that 401 mg/dL was the statistical mode for 122 participants (12.9% of the combined sample; n=40 in Lynch, n=32 in Aleppo, n=28 in Brown, n=22 in Wadwa).

**Figure 1.**
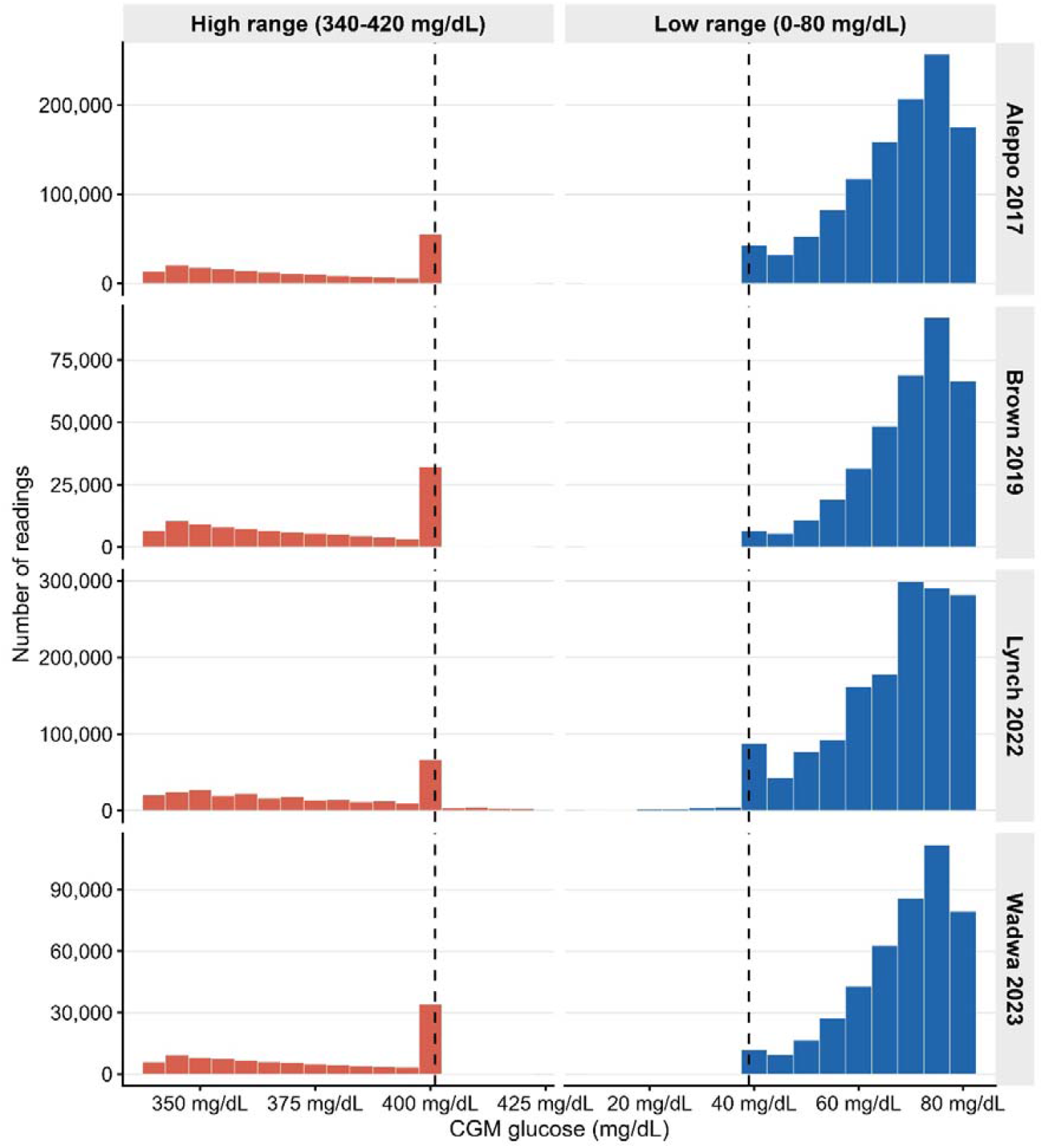
Distribution of CGM readings near sensor measurement limits across four datasets. Histograms show of glucose frequencies in the low-range window (0-80 mg/dL; left) and high-range window (340-420 mg/dL; right) for each dataset. Bin width = 5 mg/dL. Dashed lines indicate the modal cap values (39 and 401 mg/dL). The pronounced spikes at 39 and 401 mg/dL confirm systematic truncation at sensor limits.

### Upper-cap runs are frequently prolonged

In Lynch, Brown, and Aleppo, upper-cap runs were common and often sustained (Table 2, Figure 2). Median upper-cap run durations were 35, 35, and 40 minutes respectively; 36.1-38.8% of runs lasted ≥60 minutes, and over 55% lasted ≥30 minutes. During these periods, CGM records can confirm only that glucose was at or above 400 mg/dL (22.2 mmol/L), and cannot provide the true glucose value. At the participant level, the median individual accumulated 363-490 minutes of total upper-cap time, with a median longest single run of 145-170 minutes. Lower-cap runs were shorter across all datasets (median 10-20 minutes), though 7.1-14.9% lasted ≥60 minutes in three datasets. Wadwa showed a markedly different pattern: 28,403 upper-cap runs with a median duration of 5 minutes and only 1.2% lasting ≥15 minutes. We suggest that this reflects the structure of the Dexcom Clarity export for this dataset; Wadwa cap run data should therefore be interpreted with caution (Supplementary File 1), and are not included in cross-dataset summaries of run duration.

**Table 2.** Duration of consecutive CGM sensor-limit capping runs by dataset and cap type. Total runs, number of participants contributing at least one run, median and mean run duration, and proportions of runs (not participants) meeting duration thresholds (≥15 min, ≥30 min, ≥60 min). A run was defined as consecutive capped readings; duration was calculated as the interval between the first and last reading plus one 5-minute sampling interval. Wadwa 2023^10^ data should be interpreted with caution (see Supplemental file 1).

| Dataset | Cap type | N runs | N participants affected | Median duration (mins) | Mean duration (mins) | Min duration (mins) | Max duration (mins) | N runs $\geq 15$ min | N runs $\geq 30$ min | N runs $\geq 60$ min | % runs $\geq 15$ min | % runs $\geq 30$ min | % runs $\geq 60$ min |
| --- | --- | --- | --- | --- | --- | --- | --- | --- | --- | --- | --- | --- | --- |
| Aleppo 2017<sup>7</sup><br>[n=226] | Low | 7199 | 222 | 15 | 23.9 | 5 | 630 | 4007 | 1779 | 508 | 55.7 | 24.7 | 7.1 |
|  | High | 3906 | 202 | 40 | 69.4 | 5 | 1230.6 | 2898 | 2265 | 1517 | 74.2 | 58 | 38.8 |
| Brown 2019<sup>8</sup><br>[n=168] | Low | 1623 | 144 | 10 | 17.2 | 5 | 290 | 709 | 206 | 45 | 43.7 | 12.7 | 2.8 |
|  | High | 2287 | 122 | 35 | 71.4 | 5 | 1709.9 | 1778 | 1323 | 825 | 77.7 | 57.8 | 36.1 |
| Lynch 2022<sup>9</sup><br>[n=451] | Low | 14092 | 418 | 20 | 34.3 | 5 | 1469.9 | 8655 | 4975 | 2106 | 61.4 | 35.3 | 14.9 |
|  | High | 9265 | 395 | 35 | 67.8 | 5 | 3374.8 | 6861 | 5096 | 3253 | 74.1 | 55 | 35.1 |
| Wadwa 2023<sup>10</sup><br>[n=103] | Low | 7386 | 103 | 5 | 6.5 | 5 | 830 | 306 | 48 | 9 | 4.1 | 0.6 | 0.1 |
|  | High | 28403 | 95 | 5 | 5.8 | 5 | 1160 | 352 | 217 | 119 | 1.2 | 0.8 | 0.4 |

**Figure 2.**
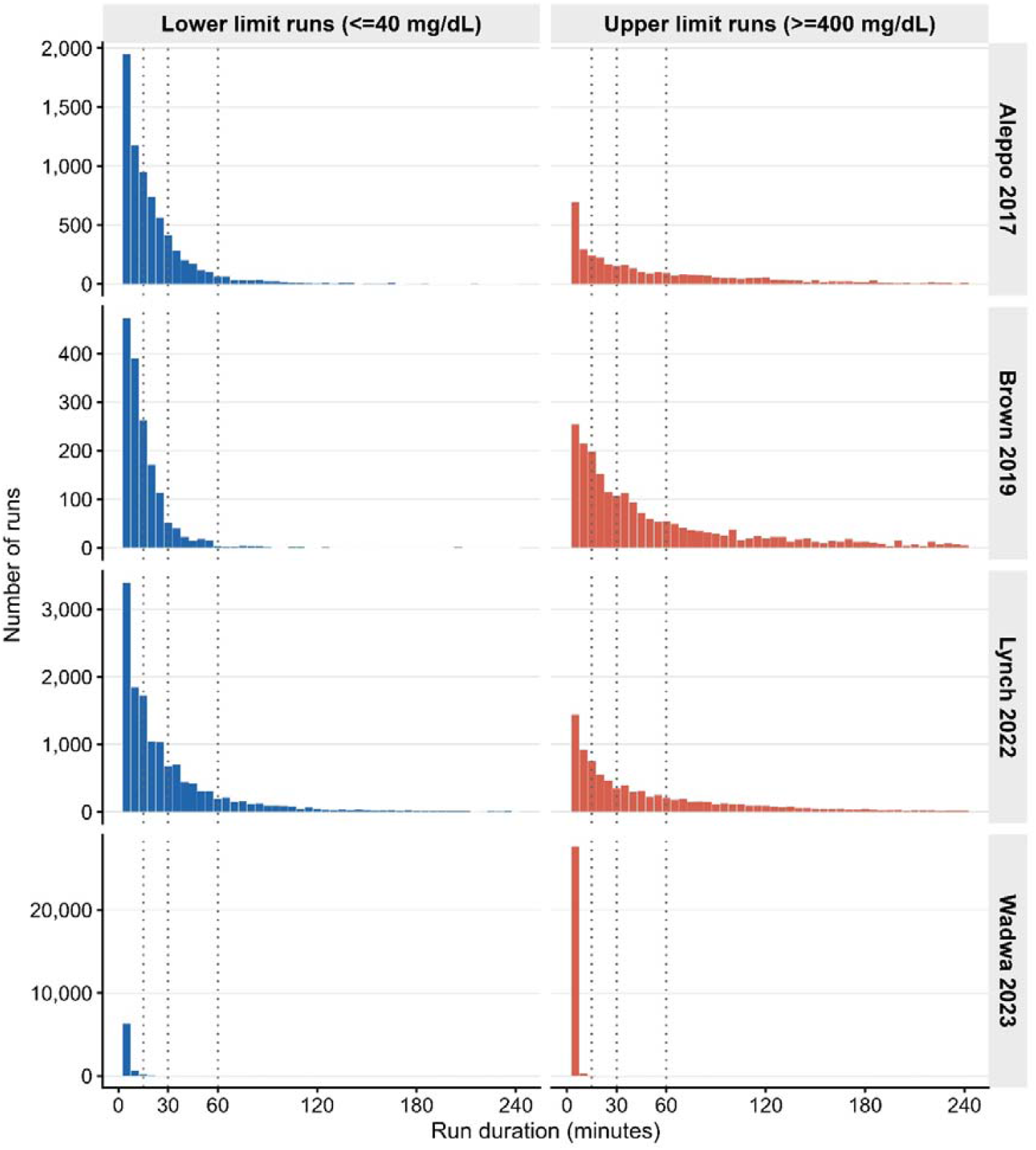
Duration of consecutive lower-limit (≤40 mg/dL) and upper-limit (≥400 mg/dL) capping runs across four datasets. Runs >240 minutes are excluded for clarity. Bin width = 5 minutes. Vertical dotted lines indicate 15-, 30-, and 60-minute thresholds.

### Sensitivity analysis: Dexcom G4 Platinum versus Dexcom G6 sensors

Upper-limit capping prevalence did not differ significantly between the Dexcom G4 dataset (Aleppo, n=226) and the three combined Dexcom G6 datasets (Brown, Lynch, Wadwa; n=722) for percentage of readings at the upper cap (median 0.116% vs 0.151%, p=0.091), total upper-cap time (388 vs 300 minutes, p=0.171), or number of upper-cap runs (7 vs 8, p=0.202) (Supplementary table S14). The longest single upper-cap run was significantly longer in the G4 dataset than in the G6 datasets (150 vs 95 minutes, p<0.001), though this most likely reflects the older age and narrower HbA1c distribution of the Aleppo cohort rather than a sensor-specific effect, since the G4’s higher measurement error (mean absolute relative difference (MARD) ∼14%) would not be expected to generate longer sustained runs at the sensor limit. Lower-limit capping burden was significantly higher in the G4 dataset across several metrics. Notably, upper-cap prevalence also differed significantly across the three G6 datasets (Kruskal-Wallis χ^2^=19.2, df=2, p<0.001), with Brown showing substantially lower median capping (0.069%) than Lynch 222 (0.174%) or Wadwa (0.177%) (Supplementary tables S15-S16), consistent with the older age and better glycaemic control of the Brown cohort rather than any sensor-specific effect. Together these findings indicate that the observed capping patterns are not attributable to sensor generation.

### Upper-limit capping shows no sex effect, but is more common in younger patients

Sex was not consistently associated with capping across these four datasets (Supplementary Table S7). In Brown, males had significantly higher upper-limit capping than females (median 0.135% vs 0.040% of readings, p=0.017), with greater total upper-cap time (223 vs 100 minutes, p=0.028) and longer maximum runs (125 vs 68 minutes, p=0.024). Conversely, in Aleppo, females had longer upper-cap runs (p=0.042). These patterns are unlikely to reflect a true biological effect. No significant sex differences in lower-limit capping were observed in any dataset.

Age showed a consistent inverse association with upper-limit capping across three datasets (Figure 3, Supplementary Tables S5-S6): Spearman ρ = −0.471 in Brown 2019, −0.378 in Lynch, and −0.202 in Aleppo (all p≤0.002). In Brown, participants under 18 had a median 0.300% of readings at the upper cap versus 0.015% in those aged 40-59 (Kruskal-Wallis p<0.001); in Lynch, 0.494% versus 0.052% (p<0.001). Full age group data are in Supplementary Table S5 and Supplementary Figure S1. No age associations were observed in Wadwa (all p>0.58), although this is to be expected as that study only enrolled participants under 18. Associations between age and lower-limit capping were less consistent: significant inverse correlations were observed in Aleppo (ρ=-0.324, p<0.001) and Brown (ρ=-0.157, p=0.042) but not in Lynch (ρ=0.024, p=0.610), where a small positive correlation with total time at the lower cap was observed (ρ=0.133, p=0.005), suggesting older participants in this dataset may experience more prolonged very low glucose events (≤40 mg/dL; ≤2.2 mmol/L) despite similar overall lower-limit capping rates.

**Figure 3.**
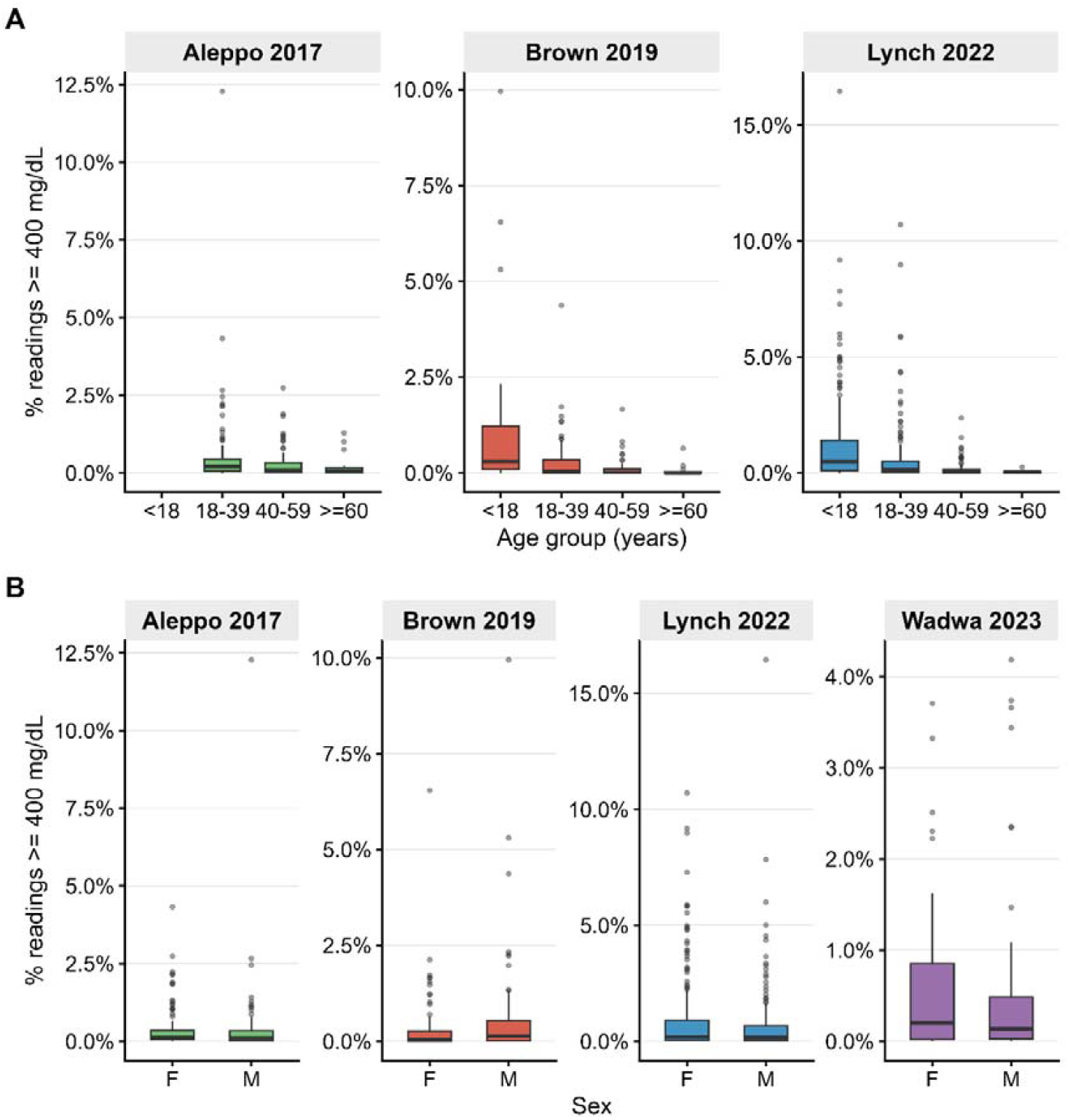
Upper-limit CGM capping by age group and sex. (A) Percentage of readings ≥400 mg/dL by age group (<18, 18-39, 40-59, ≥60 years) in three datasets (Wadwa 2023^10^ excluded; enrolled exclusively participants under 18). (B) Percentage ≥400 mg/dL by sex in all four datasets. Boxes show median and interquartile range; whiskers extend to 1.5×IQR.

### HbA1c and upper-limit capping burden

Baseline HbA1c was positively and consistently associated with upper-limit capping burden across all four datasets and all three capping metrics (all p<0.001; Supplementary Tables S11-13, Supplementary Figure S7). Across the combined cohort of 945 participants for which data were available, the Spearman correlation between HbA1c and the percentage of readings at the upper cap was ρ=0.513 (95% CI 0.465–0.559), between HbA1c and total upper-cap time was ρ=0.470 (95% CI 0.419–0.518), and between HbA1c and the longest single upper-cap run was ρ=0.375 (95% CI 0.319– 0.429). Within individual datasets, correlations were moderate to strong and most pronounced in the G6 sensor datasets: ρ ranged from 0.282 to 0.624 for percentage of readings capped, 0.389 to 0.624 for total capping time, and 0.282 to 0.601 for longest run, with the older Dexcom G4 dataset of Aleppo consistently showing the lowest correlations. Participants with HbA1c ≥9.0% accumulated substantially greater upper-limit capping burden than those with HbA1c <7.0%, consistent with the expectation that poorer overall glycaemic control produces more frequent and sustained periods of extreme hyperglycaemia beyond the sensor measurement range.

### CGM underestimates mean glucose and overestimates TIR, but not due to capping

The Aleppo trial included a CGM+BGM arm which provided contemporaneous, CGM-independent glucose measurements. We used this to first validate CGM capping events against paired BGM readings, and then to compare CGM- and BGM-derived glucose metrics across equivalent monitoring periods. Of 173,470 BGM readings from the 77 BGM+CGM participants, 145,596 (83.9%) were matched to a CGM reading within ±5 minutes (Figure 4, Supplementary Table S3). Of 682 lower-cap matched pairs, 274 (40.2%) had a paired BGM reading below 54 mg/dL (3.0 mmol/L), confirming clinically significant hypoglycaemia; 208 (30.5%) had a BGM reading in the mild hypoglycaemic range (54–69 mg/dL; 3.0–3.8 mmol/L); and 200 (29.3%) had a BGM reading in the normoglycaemic range (≥70 mg/dL; ≥3.9 mmol/L). The median paired BGM reading at the lower CGM cap was 57 mg/dL (IQR 48–73; 3.2 mmol/L, IQR 2.7–4.1).

**Figure 4.**
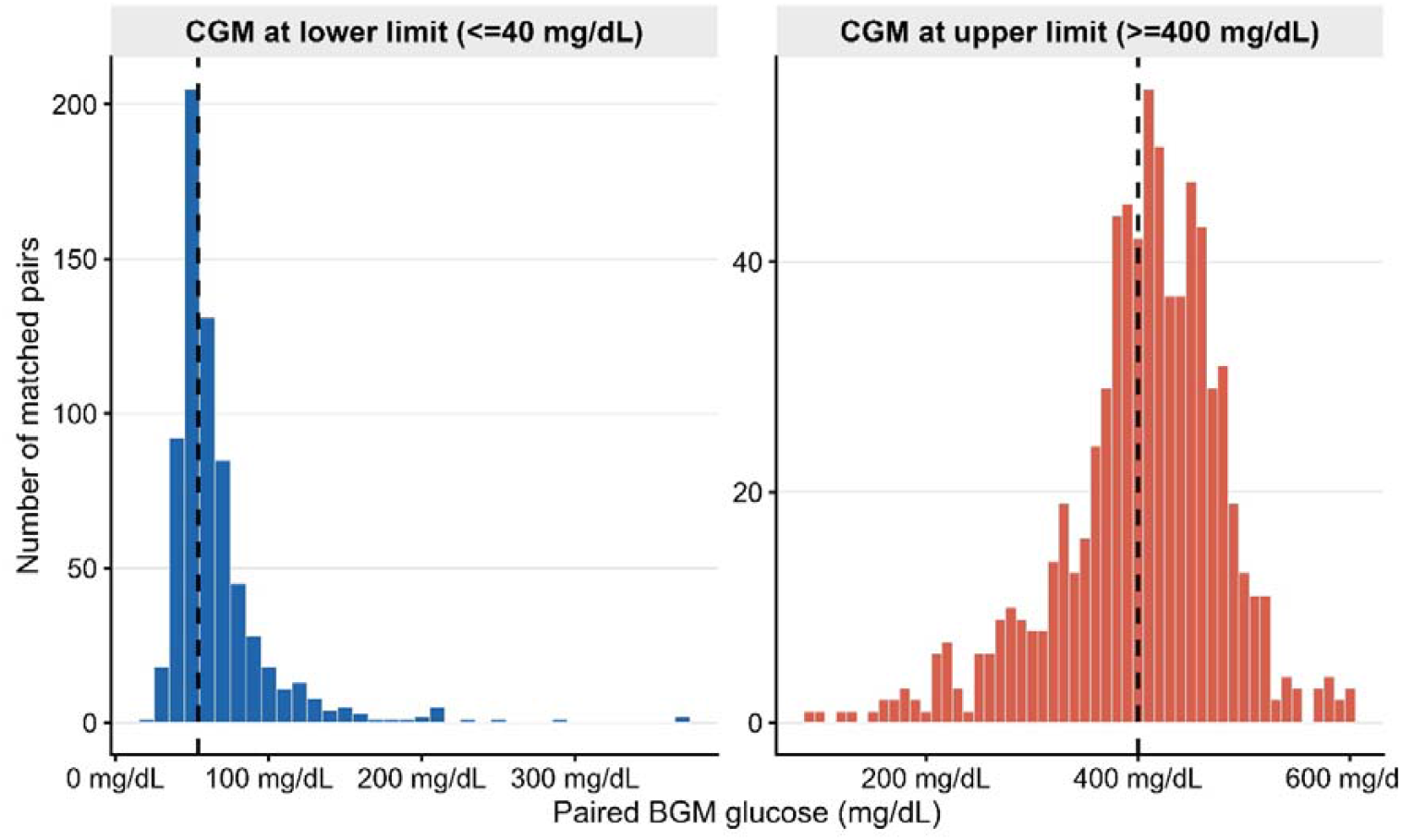
Paired BGM values at CGM sensor limits. BGM within ±5 minutes of a lower-cap (≤40 mg/dL; left panel) or upper-cap (≥400 mg/dL; right panel) CGM reading (n=77 participants). Lower limit: n=682 pairs from 71 participants; upper limit: n=738 pairs from 65 participants. Dashed lines indicate confirmation thresholds (BGM <54 mg/dL and BGM ≥400 mg/dL). Bin width = 10 mg/dL.

Of 738 upper-cap matched pairs, 436 (59.1%) had a paired BGM reading at or above 400 mg/dL (22.2 mmol/L), confirming marked hyperglycaemia; 291 (39.4%) had a BGM reading in the hyperglycaemic but sub-cap range (180–399 mg/dL; 10.0–22.2 mmol/L); and 11 (1.5%) had a BGM reading in the normoglycaemic range (<180 mg/dL; <10.0 mmol/L). The median paired BGM reading at the upper CGM cap was 411 mg/dL (IQR 369–454; 22.8 mmol/L, IQR 20.5–25.2). These matched-pair data confirm that CGM sensor caps are reached during a range of glycaemic states, from genuine hypoglycaemia to normoglycaemia at the lower limit, and from extreme to moderate hyperglycaemia at the upper limit.

We next asked whether the cumulative effect of this capping distorts CGM-derived summary metrics when calculated over a full monitoring period. Across 1,162 eligible 14-day windows, CGM systematically underestimated mean glucose relative to BGM by 10.0 mg/dL (0.6 mmol/L; CGM 158.4 vs BGM 168.4 mg/dL) and overestimated TIR by 8.3 percentage points (64.4% vs 56.1%; both p<0.001; Table 3, Figure 5, Supplementary Figure S2, Supplementary Table S8). GMI was 0.24 percentage points lower by CGM, CV 2.2 percentage points lower, and SD 7.2 mg/dL lower (all p<0.001). Time below 54 mg/dL did not differ significantly (CGM 1.0% vs BGM 1.1%, p=0.957).

**Table 3.** CGM-derived versus BGM-derived glucose metrics across 14-day monitoring windows. Full-cohort analysis (n=1,162 windows, 77 participants) and restricted analysis (≥5 BGM readings/day; n=1,008 windows, 76 participants). Values are means across windows. Difference = CGM minus BGM. All differences tested using the Wilcoxon signed-rank test, ^†^p<0.001. TBR <54 mg/dL warrants a note of caution: in our full-cohort comparison, CGM- and M-derived TBR <54 did not differ significantly (CGM 1.0% vs BGM 1.1%, p=0.957), consistent with our overall finding that capping does not distort summary metrics. A within-participant secondary analysis did identify a significant association between high lower-limit capping burden and greater CGM– M divergence (p<0.001); although, the mechanism underlying this association is not clear from the present data, based on a single dataset with 77 participants.

| Metric | All windows | | | Restricted cohort ($\geq 5$ BGM/day) | | |
| --- | --- | --- | --- | --- | --- | --- |
|  | CGM | BGM | Difference | CGM | BGM | Difference |
| Mean glucose (mg/dL) | 158.4 | 168.4 | -10.0† | 158 | 168.6 | -10.6† |
| SD (mg/dL) | 58.2 | 65.4 | -7.2† | 57.7 | 64.9 | -7.2† |
| CV (%) | 36.6 | 38.9 | -2.2† | 36.5 | 38.6 | -2.1† |
| GMI (%) | 7.1 | 7.3 | -0.2† | 7.1 | 7.3 | -0.2† |
| TIR 70–180 mg/dL (%) | 64.4 | 56.1 | 8.3† | 64.5 | 56 | 8.5† |
| TAR $> 180$ mg/dL (%) | 31.7 | 39.2 | -7.4† | 31.6 | 39.4 | -7.7† |
| TAR $> 250$ mg/dL (%) | 8.8 | 13 | -4.2† | 8.6 | 12.9 | -4.3† |
| TBR $< 54$ mg/dL (%) | 1 | 1.1 | -0.1 | 1 | 1.1 | -0.1 |

**Figure 5.**
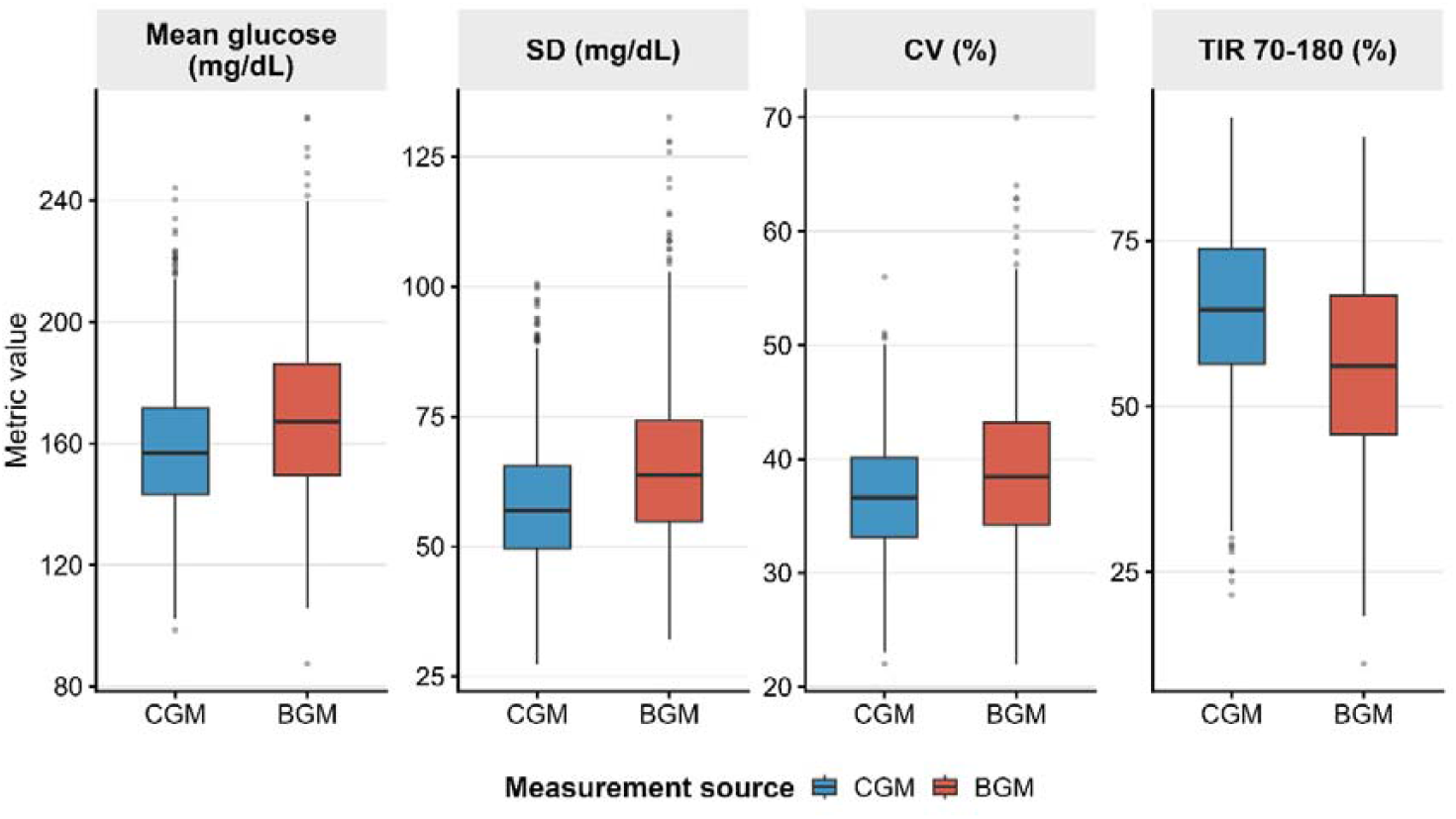
Comparison of CGM- and BGM-derived glucose metrics across 1,162 14-day monitoring windows from 77 participants. Mean glucose, SD, CV, and TIR 70–180 mg/dL were calculated separately from CGM (blue) and BGM (red) data. Boxes show median and IQR; whiskers extend to 1.5×IQR.

To test whether sensor limit capping specifically distorted CGM-derived glucose metrics (rather than simply co-occurring with high glucose) we examined whether windows with greater capping burden showed greater divergence between CGM-derived and BGM-derived metrics. If capping were the cause of the systematic CGM-BGM offset, windows in which more CGM readings were capped should show a larger gap between the two methods. We used CGM-BGM difference as the outcome rather than CGM values alone because this isolates the contribution of capping from the underlying glycaemic state: a window with genuinely high glucose would drive both CGM and BGM values upward together, whereas a capping artefact would inflate CGM-derived metrics relative to BGM. Our within-participant analysis, comparing each participant’s own higher- and lower-capping windows, provides the most rigorous version of this test, as it additionally controls for stable between-person differences in the physiological interstitial-capillary glucose offset that would otherwise confound cross-participant comparisons.

The within-participant analysis found no significant difference in metric divergence between each participant’s higher- and lower-capping windows for TIR (median Δ −0.8 percentage points, p=0.22), mean glucose (median Δ +0.6 mg/dL, p=0.90), CV (p=0.37), or SD (p=0.80). The only significant within-participant finding was for TBR <54 mg/dL (p<0.001). The systematic CGM-BGM offset in summary metrics therefore reflects the expected physiological interstitial-capillary glucose difference, not sensor capping. As a secondary analysis, between-window tertile comparisons similarly showed no monotonic dose-response between capping burden and metric divergence: median TIR divergence was 7.8, 8.4, and 6.3 percentage points across low-, mid-, and high-capping tertiles respectively, with the largest divergence in the mid-rather than the high-burden tertile, inconsistent with a causal effect (Supplementary Figures S3-S6, Tables S4, S9-S10).

## Discussion

To our knowledge, this is the first systematic characterisation of device limit-induced CGM measurement limit capping. We find such capping to be near-universal, with sustained periods during which true glucose severity cannot be assessed. Between 93.5% and 100% of participants experienced at least one capped reading, and upper-cap runs lasted a median of 35-40 minutes in the three datasets for which the duration analysis could be performed, with over one third persisting beyond 60 minutes. During these events, a clinician or algorithm examining the CGM trace knows only that glucose was at or above 400 mg/dL, not the actual figure. Measures of severe hyperglycaemia will consequently underestimate the true extent of extreme glucose excursions. The finding that 401 mg/dL was the modal CGM reading for 122 participants (12.9% of the combined sample) illustrates the practical scale of this censoring: for these individuals, the single most frequently recorded glucose value in their entire monitoring period indicated only that the sensor had reached its upper limit.

This limitation disproportionately affects younger patients. The consistent inverse association between age and upper-limit capping (Spearman ρ −0.20 to −0.47 across three independent datasets) reflects the well-established higher prevalence of severe hyperglycaemia in adolescents and young adults with type 1 diabetes^13–15^. For clinicians reviewing CGM data in younger patients and for researchers conducting paediatric or mixed-age trials, the degree to which CGM captures the true severity of extreme glucose events varies substantially by age. In trials comparing glycaemic outcomes between treatment arms, if both arms include participants with frequent readings above 400 mg/dL, treatment differences in severe hyperglycaemia may be systematically attenuated in CGM-based analyses, a bias that would be greatest in younger cohorts.

Despite this, we find that CGM capping does not substantially distort summary glucose metrics most commonly used in clinical practice and trials (mean glucose, TIR, GMI, CV, and SD) at the levels of prevalence observed here. Comparing each participant’s higher-versus lower-capping windows showed no significant effect of capping burden on any of these metrics (all p>0.2). The systematic offset between CGM and BGM (CGM underestimating mean glucose by 10 mg/dL, overestimating TIR by 8.3 percentage points, and underestimating TAR >180 mg/dL by 7.4 percentage points) is therefore attributable to the well-known physiological lag between interstitial and capillary glucose^16–18^, rather than to a capping artefact.

Olsen et al. recently proposed a Bayesian imputation model to reconstruct glucose values beyond the upper sensor limit, demonstrating good performance even at artificially imposed censoring levels of up to 30% of readings^19^. Our findings suggest that such correction may not be necessary for standard summary metrics at the capping burdens observed in these trials, where fewer than 1% of readings were capped. Whether imputation would materially alter metrics in individuals with substantially higher capping burden, such as the 122 participants for whom 401 mg/dL was the modal reading, remains an open question.

Our datasets primarily comprise Dexcom G4 and G6 systems rather than the newest CGM platforms such as Dexcom G7 or FreeStyle Libre 3. However, contemporary CGM systems continue to operate within finite measurement ranges, and therefore remain susceptible to the same underlying censoring phenomenon. The principal unresolved question is not whether capping occurs in newer systems, but whether its prevalence and duration differ materially from those observed here. Large participant-level datasets from these newer-generation sensors are not yet publicly available for independent analysis.

Our BGM comparison is restricted to a single arm of a single trial using the older Dexcom G4 Platinum sensor (mean absolute relative difference (MARD) ∼14%), which has lower accuracy than the Dexcom G6 (MARD ∼9%) used in three of our four datasets. The magnitude of the CGM–BGM offset we observed (10 mg/dL in mean glucose; 8.3 percentage points in TIR) may therefore overestimate the analogous offset for contemporary sensors. The core finding (that capping burden does not drive metric divergence) is unlikely to be sensor-specific, as it reflects glucose physiology rather than sensor accuracy; and the capping prevalence and run duration findings hold across three G6 datasets. Replication of the metric comparison in a G6, G7, or Libre 3 dataset with paired reference measurements is an important priority. Our ±5-minute matching window also may not fully account for interstitial-capillary lag^16–18^, which varies with rates and directions of glucose change. Furthermore, our within-participant analysis tests whether normal variation in capping burden across monitoring windows affects summary metrics; it does not address whether metric distortion becomes clinically meaningful in individuals with extreme capping, for example, those for whom 401 mg/dL was the modal CGM reading. This question will require further investigation in studies with larger BGM-matched samples and sufficient representation of participants with very high upper- or lower-capping prevalence.

Standard CGM reporting software does not currently flag when readings have been capped. We therefore suggest that CGM reports and trial summaries should flag the presence and burden of device-limit capping where it occurs, including the proportion of capped readings and the duration of the longest capped run (Figure 6).

**Figure 6.**
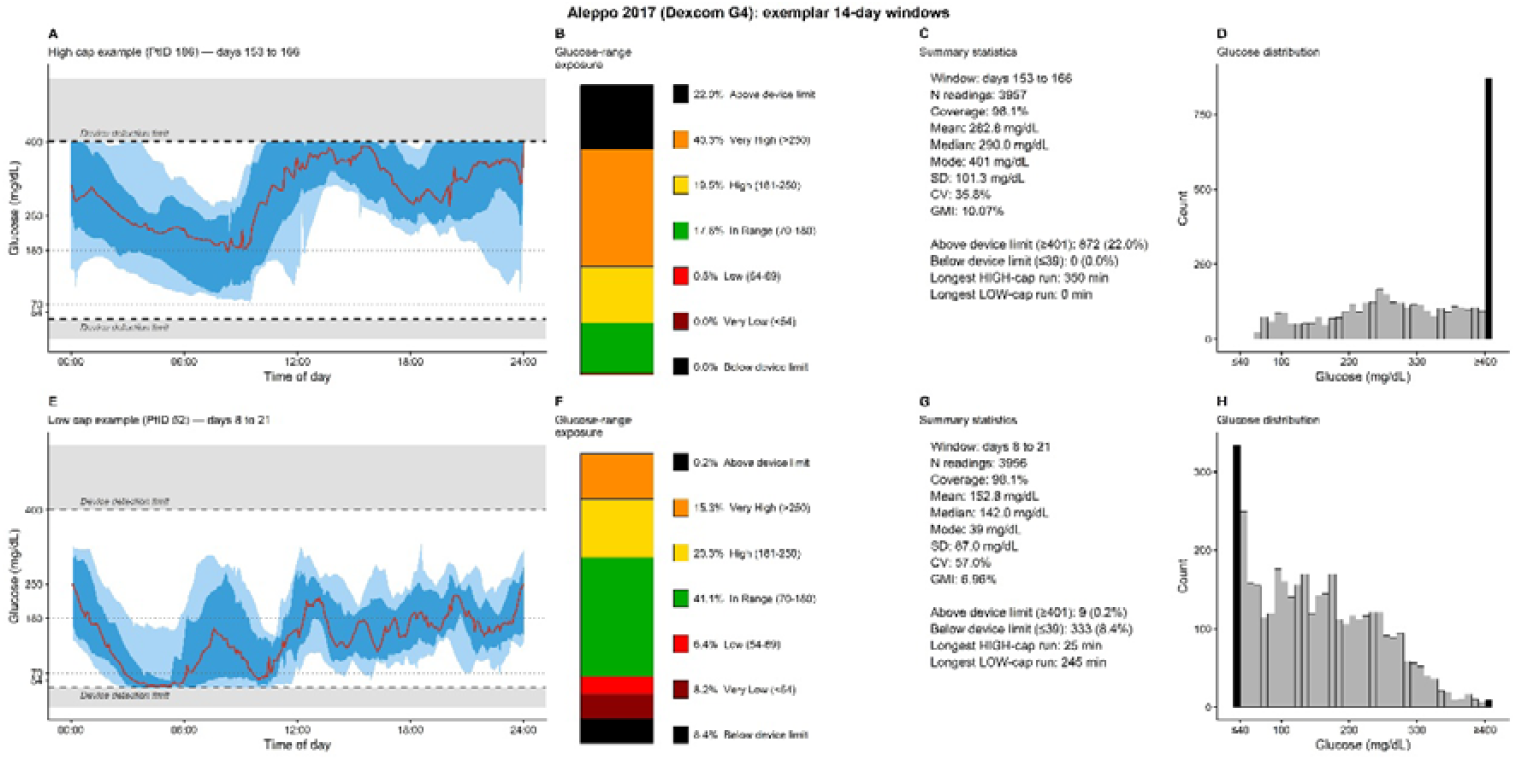
Proposed amended reporting format for CGM sensor measurement limit capping, illustrated using two exemplar 14-day windows from the Aleppo 2017^7^ Replace-BG trial (Dexcom G4 Platinum) for the participant with the highest proportion of upper-limit capped readings (A-D) and the participant with the highest proportion of lower-limit capped readings (E-H). Panels A and E show the ambulatory glucose profile (AGP) with median glucose (red line) and shaded 25–75% and 10–90% interpercentile ranges. Grey shaded bands above and below the dashed device detection limit lines (400 and 40 mg/dL) indicate the glucose range that cannot be measured by the device. Panels B and F show glucose-range exposure as a stacked bar chart, with black segments at the top and bottom representing readings at or beyond the device detection limits, extending the standard display. Panels C and G summarise conventional window-level metrics alongside device-limit capping statistics (frequency and longest run duration), and panels D and H show the glucose frequency distribution with lower and upper capped bins shaded black. These display elements are proposed as optional additions to standard CGM reporting software, to be shown only when capping is present.

In summary, CGM devices confirm that extreme glucose events have occurred but cannot quantify their severity. Capping is near-universal, disproportionately affects younger patients and those with higher HbA1c, and does not substantially distort standard summary metrics at the prevalence levels observed here. CGM reports and trial summaries should flag capping burden so that clinicians and researchers can identify those for whom this censoring is most consequential.

## Data Availability

No new data were produced

