## Supplemental tables for "Prevalence, duration, and clinical implications of Continuous Glucose Monitor (CGM) measurement limit capping in type 1 diabetes"

**Supplementary tables**

**Supplementary Table S1.** Deduplication summary by dataset. Number of fully duplicate records removed (identical participant ID, timestamp, and glucose value) and timestamp-conflicting records removed (same participant and timestamp, different glucose value). % removed = total records removed as a percentage of input records.

| **Dataset** | **Input rows** | **After exact deduplication** | **Exact duplicates removed** | **After timestamp deduplication** | **Timestamp conflicts removed** | **Total removed** | **% removed** |
| --- | --- | --- | --- | --- | --- | --- | --- |
| Aleppo 2017 | 14950661 | 14949734 | 927 | 14948968 | 766 | 1693 | 0.0113 |
| Brown 2019 | 7483946 | 7483931 | 15 | 7480842 | 3089 | 3104 | 0.0415 |
| Lynch 2022 | 18168110 | 18168097 | 13 | 18159404 | 8693 | 8706 | 0.0479 |
| Wadwa 2023 | 7232665 | 7232665 | 0 | 6401403 | 831262 | 831262 | 11.5 |
| *TOTAL* | *47835382* | *47834427* | *955* | *46990617* | *843810* | *844765* | *1.77* |

**Supplementary Table S2a.** Spike ratios at lower sensor measurement limits by dataset. Count of readings at the lower modal cap value (39 mg/dL), mean count of the two immediately adjacent 5 mg/dL bins (34–38 and 40–44 mg/dL), and spike ratios. NA indicates zero readings in that bin (strict device capping).

| **Dataset** | **Total readings** | **N readings 34–38 mg/dL** | **N readings at 39 mg/dL [%]** | **N readings 40–44 mg/dL** | **Mean adjacent bin count** | **Spike ratio vs below** | **Spike ratio vs above** | **Spike ratio vs mean** |
| --- | --- | --- | --- | --- | --- | --- | --- | --- |
| Aleppo 2017 | 14948968 | 74 | 28929 [0.1935] | 24481 | 12277.5 | 390.93 | 1.18 | 2.36 |
| Brown 2019 | 7480842 | 0 | 4875 [0.0652] | 3330 | 1665 | NA | 1.46 | 2.93 |
| Lynch 2022 | 18159404 | 7779 | 61812 [0.3404] | 39478 | 23628.5 | 7.95 | 1.57 | 2.62 |
| Wadwa 2023 | 6401403 | 0 | 8360 [0.1306] | 6567 | 3283.5 | NA | 1.27 | 2.55 |

**Supplementary Table S2b.** Spike ratios at upper sensor measurement limits by dataset. Count of readings at the upper modal cap value (401 mg/dL), mean count of the two immediately adjacent 5 mg/dL bins (396–400 and 402–406 mg/dL), and spike ratios. NA indicates zero readings in that bin (strict device capping).

| **Dataset** | **Total readings** | **N readings 396–400 mg/dL** | **N readings at 401 mg/dL [%]** | **N readings 402–406 mg/dL** | **Mean adjacent bin count** | **Spike ratio vs below** | **Spike ratio vs above** | **Spike ratio vs mean** |
| --- | --- | --- | --- | --- | --- | --- | --- | --- |
| Aleppo 2017 | 14948968 | 8290 | 48956 [0.3275] | 62 | 4176 | 5.91 | 789.61 | 11.72 |
| Brown 2019 | 7480842 | 2810 | 30608 [0.4092] | 0 | 1405 | 10.89 | NA | 21.79 |
| Lynch 2022 | 18159404 | 10715 | 57869 [0.3187] | 5248 | 7981.5 | 5.4 | 11.03 | 7.25 |
| Wadwa 2023 | 6401403 | 3094 | 32203 [0.5031] | 0 | 1547 | 10.41 | NA | 20.82 |

**Supplementary Table S3.** Per-participant paired BGM values at CGM sensor measurement limits: confirmation analysis. For each participant and cap type (lower ≤40 mg/dL; upper ≥400 mg/dL), the number of matched CGM–BGM pairs (BGM taken within ±5 minutes of a capped CGM reading), number confirmed (BGM <54 mg/dL for lower-limit events; BGM ≥400 mg/dL for upper-limit events), confirmation rate, and median BGM value at each limit. Blank cells indicate no matched pairs of that cap type for that participant; zero values indicate pairs existed but none met the confirmation threshold. BGM values in mg/dL.

| **Participant ID** | **N lower-cap pairs** | **N upper-cap pairs** | **N confirmed lower** | **N confirmed upper** | **% confirmed lower** | **% confirmed upper** | **Median BGM at lower cap** | **Median BGM at upper cap** |
| --- | --- | --- | --- | --- | --- | --- | --- | --- |
| 101 | 7 | 1 | 0 | 0 | 0 | 0 | 93 | 212 |
| 106 | 16 | 15 | 10 | 4 | 62.5 | 26.7 | 52 | 391 |
| 112 | 8 |  | 1 |  | 12.5 |  | 61 |  |
| 113 | 2 | 7 | 0 | 4 | 0 | 57.1 | 82.5 | 415 |
| 119 | 51 | 8 | 37 | 6 | 72.5 | 75 | 48 | 424 |
| 123 | 6 | 33 | 3 | 30 | 50 | 90.9 | 54.5 | 448 |
| 124 | 2 | 2 | 0 | 2 | 0 | 100 | 79.5 | 433 |
| 128 | 4 | 6 | 2 | 4 | 50 | 66.7 | 57 | 449 |
| 141 | 8 | 1 | 1 | 0 | 12.5 | 0 | 75 | 331 |
| 143 | 3 | 29 | 0 | 15 | 0 | 51.7 | 80 | 404 |
| 145 | 2 | 21 | 1 | 20 | 50 | 95.2 | 49 | 474 |
| 147 | 11 | 28 | 4 | 24 | 36.4 | 85.7 | 56 | 454 |
| 148 | 7 | 15 | 1 | 2 | 14.3 | 13.3 | 56 | 371 |
| 15 | 7 |  | 3 |  | 42.9 |  | 54 |  |
| 152 | 2 |  | 1 |  | 50 |  | 63 |  |
| 155 | 22 | 3 | 10 | 0 | 45.5 | 0 | 56 | 252 |
| 157 | 8 | 1 | 2 | 0 | 25 | 0 | 55.5 | 286 |
| 158 | 19 |  | 6 |  | 31.6 |  | 64 |  |
| 164 | 2 | 34 | 0 | 26 | 0 | 76.5 | 77 | 453.5 |
| 166 | 12 | 10 | 4 | 6 | 33.3 | 60 | 54.5 | 414 |
| 167 | 13 | 18 | 8 | 15 | 61.5 | 83.3 | 53 | 433 |
| 169 | 11 | 4 | 8 | 0 | 72.7 | 0 | 51 | 324 |
| 17 | 9 |  | 4 |  | 44.4 |  | 55 |  |
| 171 | 25 | 4 | 10 | 0 | 40 | 0 | 67 | 302 |
| 179 | 3 | 12 | 2 | 7 | 66.7 | 58.3 | 51 | 413.5 |
| 184 |  | 2 |  | 0 |  | 0 |  | 186 |
| 185 | 2 |  | 0 |  | 0 |  | 94 |  |
| 197 | 10 |  | 7 |  | 70 |  | 49 |  |
| 198 |  | 3 |  | 3 |  | 100 |  | 419 |
| 203 | 4 | 1 | 1 | 0 | 25 | 0 | 59 | 283 |
| 205 | 7 | 6 | 3 | 4 | 42.9 | 66.7 | 64 | 413 |
| 206 | 41 | 14 | 20 | 9 | 48.8 | 64.3 | 54 | 411 |
| 210 | 13 | 2 | 5 | 2 | 38.5 | 100 | 58 | 439 |
| 211 | 13 | 4 | 5 | 3 | 38.5 | 75 | 58 | 424.5 |
| 213 | 2 | 9 | 0 | 0 | 0 | 0 | 70 | 367 |
| 216 | 3 |  | 2 |  | 66.7 |  | 52 |  |
| 221 | 20 | 14 | 6 | 2 | 30 | 14.3 | 64 | 369.5 |
| 224 | 2 | 8 | 1 | 0 | 50 | 0 | 89.5 | 220.5 |
| 228 | 11 | 1 | 4 | 0 | 36.4 | 0 | 67 | 223 |
| 231 | 17 | 7 | 9 | 6 | 52.9 | 85.7 | 52 | 441 |
| 232 | 20 | 10 | 10 | 7 | 50 | 70 | 52.5 | 418 |
| 233 | 30 |  | 15 |  | 50 |  | 53.5 |  |
| 239 | 1 | 65 | 0 | 39 | 0 | 60 | 70 | 407 |
| 240 | 11 | 5 | 0 | 2 | 0 | 40 | 85 | 339 |
| 241 | 2 | 4 | 0 | 3 | 0 | 75 | 90 | 464 |
| 245 | 18 | 104 | 4 | 75 | 22.2 | 72.1 | 67 | 443 |
| 248 | 6 | 4 | 5 | 3 | 83.3 | 75 | 51.5 | 421.5 |
| 249 | 6 | 1 | 2 | 1 | 33.3 | 100 | 66 | 400 |
| 256 | 4 | 11 | 1 | 11 | 25 | 100 | 61 | 486 |
| 26 | 8 | 9 | 0 | 5 | 0 | 55.6 | 73 | 456 |
| 260 | 9 | 6 | 4 | 2 | 44.4 | 33.3 | 73 | 336.5 |
| 265 | 3 | 1 | 0 | 0 | 0 | 0 | 87 | 339 |
| 27 | 11 |  | 3 |  | 27.3 |  | 77 |  |
| 271 | 2 | 8 | 1 | 4 | 50 | 50 | 74 | 394.5 |
| 273 | 2 | 9 | 0 | 5 | 0 | 55.6 | 56 | 403 |
| 277 | 4 | 7 | 0 | 4 | 0 | 57.1 | 106.5 | 400 |
| 289 | 2 | 6 | 2 | 3 | 100 | 50 | 49.5 | 378.5 |
| 29 | 17 | 52 | 7 | 24 | 41.2 | 46.2 | 58 | 391 |
| 290 | 2 | 7 | 0 | 4 | 0 | 57.1 | 140 | 411 |
| 292 | 10 | 11 | 6 | 6 | 60 | 54.5 | 46.5 | 401 |
| 293 | 1 | 2 | 0 | 0 | 0 | 0 | 93 | 369 |
| 40 | 5 | 4 | 0 | 2 | 0 | 50 | 90 | 405.5 |
| 41 | 1 | 22 | 0 | 21 | 0 | 95.5 | 250 | 451 |
| 42 | 2 | 4 | 2 | 2 | 100 | 50 | 33 | 439 |
| 43 | 5 | 4 | 1 | 0 | 20 | 0 | 80 | 307 |
| 5 | 3 | 2 | 3 | 2 | 100 | 100 | 42 | 410.5 |
| 50 | 31 | 3 | 8 | 2 | 25.8 | 66.7 | 59 | 443 |
| 55 | 10 | 4 | 2 | 0 | 20 | 0 | 62.5 | 275 |
| 61 | 4 | 2 | 3 | 2 | 75 | 100 | 46 | 423 |
| 67 |  | 9 |  | 3 |  | 33.3 |  | 383 |
| 77 |  | 8 |  | 6 |  | 75 |  | 436 |
| 78 | 14 | 5 | 2 | 4 | 14.3 | 80 | 71.5 | 421 |
| 8 | 4 | 4 | 1 | 0 | 25 | 0 | 57 | 315.5 |
| 82 | 28 | 14 | 11 | 0 | 39.3 | 0 | 55 | 351 |
| 98 | 1 | 8 | 0 | 0 | 0 | 0 | 120 | 305 |

**Supplementary Table S4.** Within-participant effect of CGM capping burden on glucose metric divergence. For each of the 65 participants with sufficient windows to contribute both a higher-capping and a lower-capping half (split at each participant's own median capping rate), the table shows mean CGM–BGM divergence in each metric for the high-cap and low-cap window halves separately, and the delta (high-cap minus low-cap divergence). A positive delta indicates greater CGM overestimation relative to BGM in higher-capping windows. Participants are also assigned to their between-participant cap burden tertile (see Supplementary Table S10). TIR, time in range 70–180 mg/dL; CV, coefficient of variation; SD, standard deviation. All glucose values in mg/dL; divergence values are CGM minus BGM.

See separate file.

**Supplementary Table S5.** Upper-limit capping by age group. Median and interquartile range of the percentage of CGM readings at or above the upper sensor limit (≥400 mg/dL), percentage at or below the lower limit (≤40 mg/dL), total capping percentage, and total minutes at each limit, by age group (<18, 18–39, 40–59, ≥60 years). Wadwa 2023 enrolled participants aged under 18 only and so is reported in a single age group. Values are medians across participants within each age group. % values are percentage of all CGM readings.

| **Dataset** | **Age group** | **N** | **% readings ≤40 mg/dL** | **% readings ≥400 mg/dL** | **% readings capped** | **Total mins ≤40 mg/dL** | **Total mins ≥400 mg/dL** |
| --- | --- | --- | --- | --- | --- | --- | --- |
| Aleppo 2017 | 18-39 | 98 | 0.232 | 0.211 | 0.472 | 680 | 695 |
|  | 40-59 | 93 | 0.124 | 0.096 | 0.262 | 401 | 335 |
|  | ≥60 | 35 | 0.080 | 0.069 | 0.169 | 275 | 250 |
| Brown 2019 | <18 | 48 | 0.050 | 0.300 | 0.433 | 112 | 642 |
|  | 18-39 | 66 | 0.046 | 0.048 | 0.165 | 100 | 125 |
|  | 40-59 | 42 | 0.017 | 0.015 | 0.077 | 48 | 45 |
|  | ≥60 | 12 | 0.070 | 0.000 | 0.096 | 135 | 0 |
| Lynch 2022 | <18 | 221 | 0.140 | 0.494 | 0.770 | 210 | 805 |
|  | 18-39 | 132 | 0.136 | 0.138 | 0.336 | 325 | 308 |
|  | 40-59 | 79 | 0.146 | 0.052 | 0.269 | 385 | 150 |
|  | ≥60 | 19 | 0.115 | 0.046 | 0.180 | 310 | 120 |
| Wadwa 2023 | <18 | 103 | 0.090 | 0.144 | 0.327 | 220 | 505 |

**Supplementary Table S6.** Spearman correlations between participant age and CGM capping metrics. Spearman rank correlation coefficients and p-values for the association between age at enrolment and: percentage of readings at the lower cap (≤40 mg/dL), percentage at the upper cap (≥400 mg/dL), total minutes at each cap, and duration of the longest single capping run at each cap. Reported for all four datasets. Wadwa 2023 enrolled participants aged under 18 years only; the restricted age range in this dataset limits the ability to detect age-related associations. p-values <0.001 are reported as such rather than as zero. *p<0.05; **p<0.01; ***p<0.001.

| **Dataset** | **Metric** | **N** | **Spearman rho** | **p-value** |  |
| --- | --- | --- | --- | --- | --- |
| **Aleppo 2017** | Longest single run (mins), upper cap | 226 | -0.145 | 0.029 | ***** |
|  | Longest single run (mins), lower cap | 226 | -0.294 | <0.001 | ******* |
|  | % readings capped (any) | 226 | -0.303 | <0.001 | ******* |
|  | % readings at upper cap | 226 | -0.202 | 0.002 | ****** |
|  | % readings at lower cap | 226 | -0.324 | <0.001 | ******* |
|  | Total minutes at upper cap | 226 | -0.183 | 0.006 | ****** |
|  | Total minutes at lower cap | 226 | -0.295 | <0.001 | ******* |
| **Brown 2019** | Longest single run (mins), upper cap | 168 | -0.45 | <0.001 | ******* |
|  | Longest single run (mins), lower cap | 168 | -0.132 | 0.088 |  |
|  | % readings capped (any) | 168 | -0.394 | <0.001 | ******* |
|  | % readings at upper cap | 168 | -0.471 | <0.001 | ******* |
|  | % readings at lower cap | 168 | -0.157 | 0.042 | ***** |
|  | Total minutes at upper cap | 168 | -0.467 | <0.001 | ******* |
|  | Total minutes at lower cap | 168 | -0.138 | 0.074 |  |
| **Lynch 2022** | Longest single run (mins), upper cap | 451 | -0.283 | <0.001 | ******* |
|  | Longest single run (mins), lower cap | 451 | 0.072 | 0.128 |  |
|  | % readings capped (any) | 451 | -0.262 | <0.001 | ******* |
|  | % readings at upper cap | 451 | -0.378 | <0.001 | ******* |
|  | % readings at lower cap | 451 | 0.024 | 0.61 |  |
|  | Total minutes at upper cap | 451 | -0.327 | <0.001 | ******* |
|  | Total minutes at lower cap | 451 | 0.133 | 0.005 | ****** |
| **Wadwa 2023** | Longest single run (mins), upper cap | 103 | -0.035 | 0.726 |  |
|  | Longest single run (mins), lower cap | 103 | -0.037 | 0.714 |  |
|  | % readings capped (any) | 103 | 0.04 | 0.686 |  |
|  | % readings at upper cap | 103 | 0.023 | 0.817 |  |
|  | % readings at lower cap | 103 | 0.016 | 0.874 |  |
|  | Total minutes at upper cap | 103 | 0.036 | 0.715 |  |
|  | Total minutes at lower cap | 103 | -0.054 | 0.588 |  |

**Supplementary Table S7.** Mann-Whitney U tests for sex differences in CGM capping metrics. Median capping values for male and female participants and Mann-Whitney U p-values for the comparison of: percentage of readings at the lower cap (≤40 mg/dL), percentage at the upper cap (≥400 mg/dL), total minutes at each cap, and duration of the longest single capping run at each cap. Reported for all four datasets. Percentage metrics are median % of all readings; duration metrics are median minutes. *p<0.05.

| **Dataset** | **Metric** | **N (male)** | **N (female)** | **Median (male)** | **Median (female)** | **p-value** |  |
| --- | --- | --- | --- | --- | --- | --- | --- |
| **Aleppo 2017** | Longest single run (mins), upper cap | 114 | 112 | 125 | 160 | 0.042 | ***** |
|  | Longest single run (mins), lower cap | 114 | 112 | 75 | 72.5 | 0.959 |  |
|  | % readings capped (any) | 114 | 112 | 0.314 | 0.375 | 0.267 |  |
|  | % readings at upper cap | 114 | 112 | 0.113 | 0.12 | 0.136 |  |
|  | % readings at lower cap | 114 | 112 | 0.13 | 0.16 | 0.392 |  |
|  | Total minutes at upper cap | 114 | 112 | 382.5 | 420 | 0.102 |  |
|  | Total minutes at lower cap | 114 | 112 | 412 | 510 | 0.422 |  |
| **Brown 2019** | Longest single run (mins), upper cap | 84 | 84 | 125 | 67.5 | 0.024 | ***** |
|  | Longest single run (mins), lower cap | 84 | 84 | 30 | 25 | 0.5 |  |
|  | % readings capped (any) | 84 | 84 | 0.208 | 0.105 | 0.064 |  |
|  | % readings at upper cap | 84 | 84 | 0.135 | 0.04 | 0.017 | ***** |
|  | % readings at lower cap | 84 | 84 | 0.047 | 0.035 | 0.665 |  |
|  | Total minutes at upper cap | 84 | 84 | 222.5 | 100 | 0.028 | ***** |
|  | Total minutes at lower cap | 84 | 84 | 87.5 | 92.6 | 0.781 |  |
| **Lynch 2022** | Longest single run (mins), upper cap | 203 | 248 | 145 | 145 | 0.364 |  |
|  | Longest single run (mins), lower cap | 203 | 248 | 85.2 | 80 | 0.651 |  |
|  | % readings capped (any) | 203 | 248 | 0.464 | 0.481 | 0.991 |  |
|  | % readings at upper cap | 203 | 248 | 0.163 | 0.183 | 0.192 |  |
|  | % readings at lower cap | 203 | 248 | 0.144 | 0.131 | 0.519 |  |
|  | Total minutes at upper cap | 203 | 248 | 340 | 475 | 0.102 |  |
|  | Total minutes at lower cap | 203 | 248 | 290 | 292.7 | 0.932 |  |
| **Wadwa 2023** | Longest single run (mins), upper cap | 50 | 53 | 12.5 | 15 | 0.813 |  |
|  | Longest single run (mins), lower cap | 50 | 53 | 15 | 15 | 0.435 |  |
|  | % readings capped (any) | 50 | 53 | 0.278 | 0.399 | 0.642 |  |
|  | % readings at upper cap | 50 | 53 | 0.137 | 0.203 | 0.722 |  |
|  | % readings at lower cap | 50 | 53 | 0.101 | 0.074 | 0.482 |  |
|  | Total minutes at upper cap | 50 | 53 | 312.5 | 590 | 0.82 |  |
|  | Total minutes at lower cap | 50 | 53 | 217.4 | 220 | 0.937 |  |

**Supplementary Table S8.** Per-participant 14-day window summary (Aleppo 2017 CGM-plus-BGM arm). For each of the 77 participants: number of eligible 14-day windows, mean BGM readings per day, mean CGM and BGM mean glucose, mean CGM–BGM divergence in mean glucose, mean CGM and BGM time in range (TIR, 70–180 mg/dL), mean TIR divergence, mean CGM and BGM glucose management index (GMI), mean overall CGM capping percentage, and number of windows containing at least one capped reading. Diff = CGM minus BGM. pp = percentage points.

| **Participant ID** | **N windows** | **BGM/day (mean)** | **CGM mean glucose (mg/dL)** | **BGM mean glucose (mg/dL)** | **Diff mean glucose (mg/dL)** | **CGM TIR (%)** | **BGM TIR (%)** | **Diff TIR (pp)** | **CGM GMI (%)** | **BGM GMI (%)** | **Mean % CGM capped** | **N windows with any cap** |
| --- | --- | --- | --- | --- | --- | --- | --- | --- | --- | --- | --- | --- |
| 101 | 15 | 11.8 | 131.9 | 145.8 | -13.8 | 80.4 | 70.9 | 9.5 | 6.47 | 6.8 | 0.11 | 8 |
| 106 | 17 | 11.3 | 155.8 | 163.5 | -7.6 | 64.3 | 59.1 | 5.2 | 7.04 | 7.22 | 0.47 | 17 |
| 112 | 14 | 7.3 | 139.1 | 136.4 | 2.7 | 74.2 | 67.6 | 6.6 | 6.64 | 6.57 | 0.28 | 13 |
| 113 | 17 | 4.7 | 162.4 | 182.8 | -20.4 | 61.2 | 51.8 | 9.4 | 7.19 | 7.68 | 0.39 | 15 |
| 119 | 13 | 9.9 | 149 | 156.1 | -7.1 | 60.7 | 51.4 | 9.3 | 6.88 | 7.04 | 1.93 | 13 |
| 123 | 18 | 9.8 | 175.9 | 191.3 | -15.3 | 52.6 | 41.5 | 11.1 | 7.52 | 7.89 | 0.84 | 16 |
| 124 | 18 | 11.2 | 151.1 | 163.2 | -12.1 | 68.5 | 62.6 | 5.9 | 6.92 | 7.21 | 0.33 | 14 |
| 128 | 10 | 9.7 | 164.6 | 182.9 | -18.4 | 61.3 | 44.4 | 16.9 | 7.25 | 7.69 | 0.15 | 9 |
| 141 | 14 | 8.5 | 156.2 | 185.7 | -29.6 | 65.5 | 42.7 | 22.8 | 7.04 | 7.75 | 0.13 | 10 |
| 143 | 17 | 14.1 | 192.5 | 195.5 | -2.9 | 44.5 | 44.4 | 0 | 7.91 | 7.99 | 0.54 | 14 |
| 145 | 15 | 4.2 | 198.7 | 209.5 | -10.8 | 41.2 | 37.9 | 3.2 | 8.06 | 8.32 | 3.06 | 15 |
| 147 | 14 | 7.9 | 178.2 | 199 | -20.8 | 50.7 | 41.5 | 9.2 | 7.57 | 8.07 | 1.11 | 14 |
| 148 | 15 | 5.4 | 193.4 | 200.8 | -7.4 | 40.2 | 37.6 | 2.7 | 7.94 | 8.11 | 2.87 | 15 |
| 15 | 13 | 16.2 | 113.3 | 120.9 | -7.7 | 85.9 | 80 | 5.9 | 6.02 | 6.2 | 0.37 | 13 |
| 152 | 13 | 8.3 | 148.3 | 173.5 | -25.3 | 72.7 | 56.7 | 16 | 6.85 | 7.46 | 0.07 | 8 |
| 155 | 18 | 9.7 | 152.2 | 166.2 | -14 | 63.7 | 52.2 | 11.5 | 6.95 | 7.29 | 0.59 | 17 |
| 157 | 13 | 10.4 | 128.9 | 132.1 | -3.1 | 78.3 | 80.3 | -2 | 6.39 | 6.47 | 0.39 | 13 |
| 158 | 18 | 11.1 | 124.6 | 121.5 | 3.1 | 86.4 | 80.8 | 5.5 | 6.29 | 6.22 | 0.15 | 12 |
| 164 | 17 | 6.7 | 182.6 | 213.1 | -30.5 | 51.1 | 39.1 | 11.9 | 7.68 | 8.41 | 1.27 | 17 |
| 166 | 17 | 10 | 164.7 | 164.9 | -0.1 | 57.4 | 53.2 | 4.1 | 7.25 | 7.25 | 0.64 | 15 |
| 167 | 7 | 4.8 | 190.8 | 174.4 | 16.4 | 47.6 | 48.8 | -1.1 | 7.87 | 7.48 | 2.45 | 7 |
| 169 | 13 | 8.7 | 158 | 196.7 | -38.7 | 61.8 | 32.6 | 29.2 | 7.09 | 8.02 | 0.58 | 13 |
| 17 | 18 | 8.8 | 143.1 | 137.1 | 6 | 74.7 | 72.3 | 2.4 | 6.73 | 6.59 | 0.3 | 14 |
| 171 | 16 | 13 | 142.5 | 144.3 | -1.8 | 72.8 | 69.6 | 3.2 | 6.72 | 6.76 | 0.32 | 14 |
| 179 | 17 | 8 | 165.6 | 159 | 6.6 | 61.7 | 64.3 | -2.6 | 7.27 | 7.11 | 1.3 | 12 |
| 184 | 18 | 4.4 | 151.2 | 148.9 | 2.2 | 72.2 | 69.8 | 2.3 | 6.93 | 6.87 | 0.12 | 3 |
| 185 | 17 | 9.1 | 169.5 | 180.1 | -10.7 | 58.5 | 50.5 | 8 | 7.36 | 7.62 | 0.07 | 5 |
| 197 | 16 | 5.6 | 135.9 | 144.8 | -9 | 75.8 | 66.9 | 8.9 | 6.56 | 6.77 | 0.13 | 16 |
| 198 | 18 | 5.3 | 167.6 | 164.5 | 3.1 | 63.5 | 63.7 | -0.2 | 7.32 | 7.24 | 0.12 | 6 |
| 203 | 15 | 6.5 | 153.2 | 151 | 2.2 | 66.1 | 65.5 | 0.6 | 6.98 | 6.92 | 0.14 | 10 |
| 205 | 18 | 10.1 | 156.8 | 164.4 | -7.6 | 64.6 | 58.6 | 6 | 7.06 | 7.24 | 0.27 | 12 |
| 206 | 17 | 16.6 | 130.4 | 156.5 | -26.2 | 80.1 | 63.5 | 16.6 | 6.43 | 7.05 | 0.5 | 16 |
| 210 | 16 | 5.2 | 150.8 | 149.3 | 1.5 | 63.4 | 55.7 | 7.7 | 6.92 | 6.88 | 0.88 | 15 |
| 211 | 17 | 7.4 | 126.7 | 142.2 | -15.5 | 80 | 69.4 | 10.6 | 6.34 | 6.71 | 0.59 | 16 |
| 213 | 14 | 9.3 | 149.3 | 165.2 | -16 | 73.8 | 54.6 | 19.1 | 6.88 | 7.26 | 0.28 | 6 |
| 216 | 17 | 4.4 | 153.9 | 141.9 | 12 | 64.5 | 56.9 | 7.6 | 6.99 | 6.71 | 0.19 | 13 |
| 221 | 14 | 14.1 | 161.5 | 160.2 | 1.4 | 62.1 | 60 | 2.2 | 7.17 | 7.14 | 0.86 | 13 |
| 224 | 17 | 9.8 | 175.3 | 197.8 | -22.5 | 56.1 | 38.6 | 17.5 | 7.5 | 8.04 | 0.18 | 7 |
| 228 | 19 | 8.9 | 132.8 | 140.2 | -7.4 | 77.8 | 74.3 | 3.5 | 6.49 | 6.66 | 0.61 | 16 |
| 231 | 14 | 9.5 | 131.7 | 150.1 | -18.4 | 77.4 | 51.6 | 25.8 | 6.46 | 6.9 | 0.35 | 13 |
| 232 | 18 | 6.7 | 151.1 | 175.9 | -24.8 | 66.3 | 40.6 | 25.7 | 6.93 | 7.52 | 0.43 | 17 |
| 233 | 18 | 11.3 | 119.1 | 121.7 | -2.5 | 73.4 | 76.1 | -2.6 | 6.16 | 6.22 | 0.6 | 18 |
| 239 | 17 | 7.3 | 213.9 | 220.5 | -6.6 | 35 | 33.6 | 1.4 | 8.43 | 8.58 | 1.45 | 14 |
| 240 | 18 | 10.7 | 145.9 | 158.7 | -12.8 | 74.3 | 67 | 7.3 | 6.8 | 7.11 | 0.65 | 17 |
| 241 | 13 | 8.9 | 168.5 | 184.2 | -15.6 | 59.7 | 46.2 | 13.6 | 7.34 | 7.71 | 0.05 | 5 |
| 245 | 17 | 9.5 | 195.6 | 211.1 | -15.5 | 45.6 | 40 | 5.5 | 7.99 | 8.36 | 3.12 | 17 |
| 248 | 14 | 6.2 | 177.8 | 175.4 | 2.5 | 52.1 | 50.1 | 1.9 | 7.56 | 7.5 | 1.04 | 14 |
| 249 | 6 | 6.4 | 154.3 | 171.5 | -17.1 | 66 | 52 | 14 | 7 | 7.41 | 0.46 | 5 |
| 256 | 14 | 7.8 | 148.3 | 173.7 | -25.4 | 70.1 | 52.2 | 17.9 | 6.86 | 7.46 | 0.54 | 14 |
| 26 | 18 | 9.7 | 162.9 | 178.9 | -16 | 64.1 | 53.5 | 10.7 | 7.21 | 7.59 | 0.13 | 9 |
| 260 | 18 | 5.2 | 155 | 159 | -4.1 | 65.5 | 59 | 6.5 | 7.02 | 7.11 | 0.66 | 14 |
| 265 | 17 | 4 | 139 | 154.9 | -15.9 | 82.6 | 69.9 | 12.7 | 6.63 | 7.01 | 0.11 | 7 |
| 27 | 17 | 4.3 | 142.3 | 147.7 | -5.4 | 69.3 | 66.1 | 3.2 | 6.71 | 6.84 | 0.48 | 16 |
| 271 | 13 | 8 | 159.7 | 167.8 | -8.1 | 67.3 | 62.1 | 5.3 | 7.13 | 7.32 | 0.21 | 8 |
| 272 | 14 | 8.6 | 149.3 | 150.1 | -0.8 | 80.2 | 79.5 | 0.6 | 6.88 | 6.9 | 0.01 | 1 |
| 273 | 17 | 9.6 | 168.7 | 178.5 | -9.8 | 58.1 | 51.1 | 7 | 7.35 | 7.58 | 0.19 | 9 |
| 277 | 16 | 7.9 | 168.7 | 168.9 | -0.2 | 61.2 | 60.5 | 0.7 | 7.35 | 7.35 | 0.18 | 9 |
| 289 | 4 | 4.2 | 199.7 | 190.4 | 9.3 | 39.8 | 38.9 | 0.9 | 8.09 | 7.86 | 2.92 | 4 |
| 29 | 18 | 12 | 187.1 | 193.7 | -6.6 | 48.7 | 43.1 | 5.6 | 7.79 | 7.94 | 1.38 | 18 |
| 290 | 15 | 4.4 | 167.6 | 181 | -13.4 | 61.5 | 52.7 | 8.8 | 7.32 | 7.64 | 0.82 | 13 |
| 292 | 12 | 10.9 | 140.1 | 141.2 | -1.2 | 76.6 | 70.4 | 6.2 | 6.66 | 6.69 | 0.17 | 9 |
| 293 | 13 | 6.6 | 177.8 | 212 | -34.2 | 54.2 | 31.8 | 22.4 | 7.56 | 8.38 | 0.06 | 5 |
| 40 | 16 | 8.5 | 159 | 173.4 | -14.4 | 67.3 | 56.7 | 10.6 | 7.11 | 7.46 | 0.15 | 6 |
| 41 | 13 | 8.5 | 182.8 | 199.8 | -17 | 53.1 | 43.4 | 9.7 | 7.68 | 8.09 | 0.4 | 10 |
| 42 | 14 | 7.9 | 156 | 153.3 | 2.8 | 65.9 | 59.8 | 6.1 | 7.04 | 6.98 | 0.25 | 11 |
| 43 | 13 | 10.6 | 147.6 | 165 | -17.4 | 69.8 | 59.1 | 10.7 | 6.84 | 7.26 | 0.2 | 9 |
| 5 | 10 | 7.6 | 149.6 | 173.3 | -23.7 | 72.5 | 53 | 19.5 | 6.89 | 7.46 | 0.13 | 6 |
| 50 | 13 | 10 | 150.2 | 181.8 | -31.6 | 67 | 42.3 | 24.8 | 6.9 | 7.66 | 0.49 | 13 |
| 55 | 9 | 14.4 | 132.9 | 152.3 | -19.4 | 82.3 | 67.8 | 14.5 | 6.49 | 6.95 | 0.25 | 8 |
| 61 | 17 | 8.9 | 163.2 | 166 | -2.8 | 60.2 | 59.2 | 1 | 7.21 | 7.28 | 0.36 | 13 |
| 67 | 14 | 7.6 | 186.8 | 196.2 | -9.4 | 49.3 | 42.9 | 6.5 | 7.78 | 8 | 0.5 | 11 |
| 77 | 18 | 9.4 | 165.9 | 182.8 | -16.8 | 62.4 | 50.5 | 11.8 | 7.28 | 7.68 | 0.22 | 10 |
| 78 | 14 | 7.7 | 162.5 | 179.8 | -17.4 | 62.3 | 51.5 | 10.8 | 7.2 | 7.61 | 0.41 | 13 |
| 8 | 13 | 5.9 | 160.7 | 171 | -10.2 | 63.8 | 54.1 | 9.7 | 7.15 | 7.4 | 0.22 | 12 |
| 82 | 14 | 12.9 | 180.9 | 181.6 | -0.7 | 49.3 | 47.3 | 2.1 | 7.64 | 7.65 | 0.74 | 14 |
| 9 | 18 | 10.4 | 154.4 | 158.5 | -4.1 | 70.9 | 67.6 | 3.3 | 7 | 7.1 | 0.04 | 2 |
| 98 | 15 | 10 | 174.6 | 185.5 | -10.9 | 55.5 | 46.4 | 9.1 | 7.49 | 7.75 | 0.07 | 5 |

**Supplementary Table S9.** Between-participant cap burden tertile classification and metric divergence summary. Participants assigned to tertiles based on mean capping percentage across all eligible 14-day windows: T1 (low burden), T2 (mid burden), T3 (high burden). Values shown are median (IQR) CGM–BGM divergence for each metric across all windows within each tertile. Diff = CGM minus BGM. pp = percentage points. TIR = time in range 70–180 mg/dL; TBR = time below range; TAR = time above range; GMI = glucose management indicator; CV = coefficient of variation; SD = standard deviation.

| **Tertile** | **N participants** | **N windows** | **Mean capping (%)** | **Mean glucose diff (pp)** | **GMI diff (pp)** | **CV diff (pp)** | **SD diff (mg/dL)** | **TIR diff (pp)** | **TBR <54 diff (pp)** | **TAR >180 diff (pp)** |
| --- | --- | --- | --- | --- | --- | --- | --- | --- | --- | --- |
| Low burden (T1) | 26 | 393 | 0.125 | -9.4 (-18.8 to 0.2) | -0.22 (-0.45 to 0.00) | -1.7 (-4.5 to 0.2) | -6.1 (-10.8 to -2.1) | 8.0 (3.0 to 14.3) | 0.00 (-0.40 to 0.20) | -7.7 (-13.6 to -1.6) |
| Mid burden (T2) | 26 | 386 | 0.377 | -10.9 (-19.6 to -2.5) | -0.26 (-0.47 to -0.06) | -2.0 (-5.0 to 1.1) | -6.8 (-12.2 to -1.9) | 8.7 (3.1 to 15.0) | 0.10 (-0.70 to 0.60) | -7.5 (-15.1 to -2.1) |
| High burden (T3) | 25 | 383 | 1.223 | -8.9 (-16.0 to 0.8) | -0.21 (-0.38 to 0.02) | -2.0 (-5.0 to 1.0) | -7.7 (-13.6 to -1.1) | 5.9 (0.4 to 11.3) | 0.20 (-0.70 to 0.85) | -5.0 (-10.3 to -0.1) |

**Supplementary Table S10.** Per-participant CGM capping burden and glucose metric divergence. One row per participant (n=77) with three or more eligible 14-day monitoring windows. Columns show mean and median overall capping percentage, mean lower-limit and upper-limit capping percentages, mean CGM-derived and BGM-derived glucose metrics averaged across all eligible windows, mean CGM–BGM divergence for each metric, and cap burden tertile assignment. Tertile boundaries based on mean overall capping percentage: T1 (low burden) <0.18%, T2 (mid burden) 0.18–0.54%, T3 (high burden) >0.54%. TIR = time in range 70–180 mg/dL; CV = coefficient of variation; SD = standard deviation. All glucose values in mg/dL; divergence values are CGM minus BGM.

| **ID** | **N windows** | **Mean % capped** | **Median % capped** | **Mean % cap low** | **Mean % cap high** | **Mean CGM TIR** | **Mean BGM TIR** | **Mean diff TIR** | **Mean CGM mean** | **Mean BGM mean** | **Mean diff mean** | **Mean CGM CV** | **Mean BGM CV** | **Mean diff CV** | **Mean CGM SD** | **Mean BGM SD** | **Mean diff SD** | **Cap tertile** |
| --- | --- | --- | --- | --- | --- | --- | --- | --- | --- | --- | --- | --- | --- | --- | --- | --- | --- | --- |
| 101 | 15 | 0.107 | 0.13 | 0.107 | 0 | 80.4 | 70.9 | 9.5 | 131.9 | 145.8 | -13.8 | 33.1 | 35.4 | -2.3 | 43.7 | 51.7 | -8 | Low burden (T1) |
| 106 | 17 | 0.472 | 0.4 | 0.271 | 0.202 | 64.3 | 59.1 | 5.2 | 155.8 | 163.5 | -7.6 | 40.3 | 43.4 | -3.1 | 62.7 | 70.8 | -8.1 | Mid burden (T2) |
| 112 | 14 | 0.282 | 0.22 | 0.282 | 0 | 74.2 | 67.6 | 6.6 | 139.1 | 136.4 | 2.7 | 35.3 | 41.3 | -6 | 49 | 56.3 | -7.2 | Mid burden (T2) |
| 113 | 17 | 0.391 | 0.18 | 0.179 | 0.212 | 61.2 | 51.8 | 9.4 | 162.4 | 182.8 | -20.4 | 39.5 | 37.4 | 2.1 | 64.1 | 68.6 | -4.4 | Mid burden (T2) |
| 119 | 13 | 1.935 | 1.48 | 1.562 | 0.374 | 60.7 | 51.4 | 9.3 | 149 | 156.1 | -7.1 | 46 | 51.1 | -5.1 | 68.6 | 79.7 | -11.1 | High burden (T3) |
| 123 | 18 | 0.843 | 0.3 | 0.181 | 0.662 | 52.6 | 41.5 | 11.1 | 175.9 | 191.3 | -15.3 | 38.5 | 39.4 | -1 | 67.6 | 75.1 | -7.5 | High burden (T3) |
| 124 | 18 | 0.328 | 0.235 | 0.28 | 0.048 | 68.5 | 62.6 | 5.9 | 151.1 | 163.2 | -12.1 | 36.1 | 33.3 | 2.8 | 54.6 | 54.3 | 0.2 | Mid burden (T2) |
| 128 | 10 | 0.154 | 0.05 | 0.046 | 0.108 | 61.3 | 44.4 | 16.9 | 164.6 | 182.9 | -18.4 | 34.1 | 39.2 | -5.1 | 56.1 | 71.3 | -15.2 | Low burden (T1) |
| 141 | 14 | 0.129 | 0.095 | 0.102 | 0.026 | 65.5 | 42.7 | 22.8 | 156.2 | 185.7 | -29.6 | 35 | 36.5 | -1.5 | 54.6 | 67.4 | -12.8 | Low burden (T1) |
| 143 | 17 | 0.544 | 0.4 | 0.054 | 0.491 | 44.5 | 44.4 | 0 | 192.5 | 195.5 | -2.9 | 35.5 | 35.9 | -0.4 | 68 | 70 | -2 | Mid burden (T2) |
| 145 | 15 | 3.065 | 2.59 | 0.289 | 2.775 | 41.2 | 37.9 | 3.2 | 198.7 | 209.5 | -10.8 | 42.3 | 44 | -1.7 | 84.1 | 92.3 | -8.2 | High burden (T3) |
| 147 | 14 | 1.111 | 0.735 | 0.249 | 0.863 | 50.7 | 41.5 | 9.2 | 178.2 | 199 | -20.8 | 44 | 44.4 | -0.4 | 78.2 | 88.2 | -9.9 | High burden (T3) |
| 148 | 15 | 2.865 | 2.37 | 1.044 | 1.821 | 40.2 | 37.6 | 2.7 | 193.4 | 200.8 | -7.4 | 43.5 | 39.2 | 4.3 | 83.9 | 78.5 | 5.4 | High burden (T3) |
| 15 | 13 | 0.368 | 0.4 | 0.368 | 0 | 85.9 | 80 | 5.9 | 113.3 | 120.9 | -7.7 | 32 | 35.9 | -4 | 36.3 | 43.7 | -7.4 | Mid burden (T2) |
| 152 | 13 | 0.067 | 0.03 | 0.067 | 0 | 72.7 | 56.7 | 16 | 148.3 | 173.5 | -25.3 | 34.5 | 37.5 | -3 | 51.1 | 65 | -13.9 | Low burden (T1) |
| 155 | 18 | 0.59 | 0.465 | 0.456 | 0.135 | 63.7 | 52.2 | 11.5 | 152.2 | 166.2 | -14 | 38.4 | 38.8 | -0.4 | 58.4 | 64.4 | -6 | High burden (T3) |
| 157 | 13 | 0.387 | 0.38 | 0.332 | 0.054 | 78.3 | 80.3 | -2 | 128.9 | 132.1 | -3.1 | 36.8 | 35 | 1.9 | 47.7 | 46.2 | 1.4 | Mid burden (T2) |
| 158 | 18 | 0.148 | 0.05 | 0.148 | 0 | 86.4 | 80.8 | 5.5 | 124.6 | 121.5 | 3.1 | 30.6 | 34.8 | -4.2 | 38.1 | 42.3 | -4.1 | Low burden (T1) |
| 164 | 17 | 1.268 | 1.28 | 0.331 | 0.937 | 51.1 | 39.1 | 11.9 | 182.6 | 213.1 | -30.5 | 42.2 | 44.5 | -2.2 | 77.1 | 94.8 | -17.7 | High burden (T3) |
| 166 | 17 | 0.639 | 0.25 | 0.235 | 0.404 | 57.4 | 53.2 | 4.1 | 164.7 | 164.9 | -0.1 | 39.2 | 43.3 | -4.1 | 64.6 | 71.2 | -6.6 | High burden (T3) |
| 167 | 7 | 2.453 | 1.4 | 0.527 | 1.927 | 47.6 | 48.8 | -1.1 | 190.8 | 174.4 | 16.4 | 44.5 | 55.5 | -11 | 84.4 | 96.2 | -11.9 | High burden (T3) |
| 169 | 13 | 0.581 | 0.5 | 0.518 | 0.063 | 61.8 | 32.6 | 29.2 | 158 | 196.7 | -38.7 | 38.1 | 36.5 | 1.6 | 60.2 | 71.7 | -11.5 | High burden (T3) |
| 17 | 18 | 0.301 | 0.305 | 0.117 | 0.184 | 74.7 | 72.3 | 2.4 | 143.1 | 137.1 | 6 | 39.6 | 42.2 | -2.7 | 56.9 | 58.2 | -1.3 | Mid burden (T2) |
| 171 | 16 | 0.318 | 0.185 | 0.279 | 0.039 | 72.8 | 69.6 | 3.2 | 142.5 | 144.3 | -1.8 | 35.4 | 40 | -4.6 | 50.5 | 57.8 | -7.3 | Mid burden (T2) |
| 179 | 17 | 1.303 | 1.02 | 0.062 | 1.242 | 61.7 | 64.3 | -2.6 | 165.6 | 159 | 6.6 | 41.5 | 42.2 | -0.8 | 68.7 | 67 | 1.7 | High burden (T3) |
| 184 | 18 | 0.116 | 0 | 0.001 | 0.114 | 72.2 | 69.8 | 2.3 | 151.2 | 148.9 | 2.2 | 32.6 | 37.8 | -5.2 | 49.4 | 56.4 | -7 | Low burden (T1) |
| 185 | 17 | 0.065 | 0 | 0.043 | 0.022 | 58.5 | 50.5 | 8 | 169.5 | 180.1 | -10.7 | 29.7 | 29.1 | 0.6 | 50.3 | 52.4 | -2.1 | Low burden (T1) |
| 197 | 16 | 0.131 | 0.09 | 0.131 | 0 | 75.8 | 66.9 | 8.9 | 135.9 | 144.8 | -9 | 37.9 | 44.4 | -6.6 | 51.5 | 64.8 | -13.3 | Low burden (T1) |
| 198 | 18 | 0.121 | 0 | 0.032 | 0.089 | 63.5 | 63.7 | -0.2 | 167.6 | 164.5 | 3.1 | 32.7 | 34.1 | -1.4 | 54.8 | 56.1 | -1.2 | Low burden (T1) |
| 203 | 15 | 0.145 | 0.1 | 0.129 | 0.015 | 66.1 | 65.5 | 0.6 | 153.2 | 151 | 2.2 | 37.3 | 38.1 | -0.8 | 56.8 | 57.2 | -0.5 | Low burden (T1) |
| 205 | 18 | 0.268 | 0.08 | 0.152 | 0.116 | 64.6 | 58.6 | 6 | 156.8 | 164.4 | -7.6 | 37 | 38.7 | -1.7 | 58 | 63.6 | -5.6 | Mid burden (T2) |
| 206 | 17 | 0.5 | 0.31 | 0.317 | 0.183 | 80.1 | 63.5 | 16.6 | 130.4 | 156.5 | -26.2 | 34.3 | 35.9 | -1.6 | 45 | 56.2 | -11.2 | Mid burden (T2) |
| 210 | 16 | 0.875 | 0.28 | 0.707 | 0.169 | 63.4 | 55.7 | 7.7 | 150.8 | 149.3 | 1.5 | 41.3 | 47.7 | -6.3 | 62.5 | 71.3 | -8.8 | High burden (T3) |
| 211 | 17 | 0.594 | 0.48 | 0.5 | 0.094 | 80 | 69.4 | 10.6 | 126.7 | 142.2 | -15.5 | 38.1 | 42 | -3.9 | 48.6 | 60.1 | -11.5 | High burden (T3) |
| 213 | 14 | 0.282 | 0 | 0.032 | 0.251 | 73.8 | 54.6 | 19.1 | 149.3 | 165.2 | -16 | 34 | 32.5 | 1.5 | 50.8 | 53.7 | -2.9 | Mid burden (T2) |
| 216 | 17 | 0.192 | 0.15 | 0.192 | 0 | 64.5 | 56.9 | 7.6 | 153.9 | 141.9 | 12 | 36.7 | 45.7 | -8.9 | 56.6 | 64.6 | -8 | Low burden (T1) |
| 221 | 14 | 0.856 | 0.635 | 0.428 | 0.427 | 62.1 | 60 | 2.2 | 161.5 | 160.2 | 1.4 | 41.5 | 42.2 | -0.7 | 66.9 | 67.5 | -0.5 | High burden (T3) |
| 224 | 17 | 0.176 | 0 | 0.022 | 0.155 | 56.1 | 38.6 | 17.5 | 175.3 | 197.8 | -22.5 | 29.3 | 28.5 | 0.8 | 51.3 | 56.1 | -4.8 | Low burden (T1) |
| 228 | 19 | 0.606 | 0.33 | 0.557 | 0.049 | 77.8 | 74.3 | 3.5 | 132.8 | 140.2 | -7.4 | 35.8 | 34.9 | 0.9 | 47.6 | 48.9 | -1.3 | High burden (T3) |
| 231 | 14 | 0.35 | 0.255 | 0.256 | 0.094 | 77.4 | 51.6 | 25.8 | 131.7 | 150.1 | -18.4 | 36.8 | 46.2 | -9.4 | 48.5 | 69.3 | -20.8 | Mid burden (T2) |
| 232 | 18 | 0.43 | 0.25 | 0.249 | 0.181 | 66.3 | 40.6 | 25.7 | 151.1 | 175.9 | -24.8 | 38.8 | 47.5 | -8.8 | 58.5 | 83.1 | -24.6 | Mid burden (T2) |
| 233 | 18 | 0.598 | 0.39 | 0.598 | 0 | 73.4 | 76.1 | -2.6 | 119.1 | 121.7 | -2.5 | 41.3 | 42 | -0.7 | 49.2 | 51.2 | -2 | High burden (T3) |
| 239 | 17 | 1.452 | 1.1 | 0.037 | 1.415 | 35 | 33.6 | 1.4 | 213.9 | 220.5 | -6.6 | 33.8 | 39.1 | -5.4 | 72.3 | 86.2 | -13.9 | High burden (T3) |
| 240 | 18 | 0.649 | 0.435 | 0.423 | 0.226 | 74.3 | 67 | 7.3 | 145.9 | 158.7 | -12.8 | 35.9 | 33.9 | 1.9 | 52.7 | 53.9 | -1.2 | High burden (T3) |
| 241 | 13 | 0.051 | 0 | 0.016 | 0.036 | 59.7 | 46.2 | 13.6 | 168.5 | 184.2 | -15.6 | 34.9 | 39.5 | -4.6 | 58.7 | 72.3 | -13.6 | Low burden (T1) |
| 245 | 17 | 3.118 | 2.63 | 0.199 | 2.918 | 45.6 | 40 | 5.5 | 195.6 | 211.1 | -15.5 | 43.5 | 47.1 | -3.6 | 84.5 | 98.6 | -14.1 | High burden (T3) |
| 248 | 14 | 1.039 | 0.675 | 0.433 | 0.604 | 52.1 | 50.1 | 1.9 | 177.8 | 175.4 | 2.5 | 41 | 45.5 | -4.5 | 72.5 | 79.4 | -6.9 | High burden (T3) |
| 249 | 6 | 0.463 | 0.45 | 0.315 | 0.147 | 66 | 52 | 14 | 154.3 | 171.5 | -17.1 | 39 | 43.2 | -4.2 | 60.5 | 74.2 | -13.7 | Mid burden (T2) |
| 256 | 14 | 0.543 | 0.44 | 0.165 | 0.377 | 70.1 | 52.2 | 17.9 | 148.3 | 173.7 | -25.4 | 40 | 43 | -3 | 59.4 | 74.8 | -15.4 | Mid burden (T2) |
| 26 | 18 | 0.131 | 0.015 | 0.036 | 0.094 | 64.1 | 53.5 | 10.7 | 162.9 | 178.9 | -16 | 34.2 | 33.6 | 0.6 | 55.6 | 60.1 | -4.5 | Low burden (T1) |
| 260 | 18 | 0.662 | 0.285 | 0.308 | 0.354 | 65.5 | 59 | 6.5 | 155 | 159 | -4.1 | 38.8 | 45.2 | -6.4 | 60.1 | 71.8 | -11.7 | High burden (T3) |
| 265 | 17 | 0.105 | 0 | 0.096 | 0.009 | 82.6 | 69.9 | 12.7 | 139 | 154.9 | -15.9 | 29 | 32.3 | -3.3 | 40.2 | 50.1 | -9.9 | Low burden (T1) |
| 27 | 17 | 0.484 | 0.41 | 0.37 | 0.114 | 69.3 | 66.1 | 3.2 | 142.3 | 147.7 | -5.4 | 39.6 | 40 | -0.5 | 56.5 | 59.3 | -2.8 | Mid burden (T2) |
| 271 | 13 | 0.205 | 0.08 | 0.162 | 0.043 | 67.3 | 62.1 | 5.3 | 159.7 | 167.8 | -8.1 | 32.2 | 30.5 | 1.7 | 51.4 | 51.2 | 0.2 | Low burden (T1) |
| 272 | 14 | 0.011 | 0 | 0.011 | 0 | 80.2 | 79.5 | 0.6 | 149.3 | 150.1 | -0.8 | 25.2 | 27.4 | -2.2 | 37.6 | 41 | -3.5 | Low burden (T1) |
| 273 | 17 | 0.189 | 0.03 | 0.005 | 0.184 | 58.1 | 51.1 | 7 | 168.7 | 178.5 | -9.8 | 36.1 | 37.3 | -1.2 | 60.8 | 66.3 | -5.5 | Low burden (T1) |
| 277 | 16 | 0.183 | 0.13 | 0.087 | 0.097 | 61.2 | 60.5 | 0.7 | 168.7 | 168.9 | -0.2 | 33.8 | 36.6 | -2.8 | 57.1 | 61.9 | -4.8 | Low burden (T1) |
| 289 | 4 | 2.915 | 2.695 | 0.188 | 2.728 | 39.8 | 38.9 | 0.9 | 199.7 | 190.4 | 9.3 | 41.2 | 47.7 | -6.5 | 81.9 | 90.2 | -8.4 | High burden (T3) |
| 29 | 18 | 1.384 | 1.095 | 0.324 | 1.061 | 48.7 | 43.1 | 5.6 | 187.1 | 193.7 | -6.6 | 39.9 | 42.4 | -2.5 | 74.6 | 82.1 | -7.5 | High burden (T3) |
| 290 | 15 | 0.823 | 0.59 | 0.128 | 0.695 | 61.5 | 52.7 | 8.8 | 167.6 | 181 | -13.4 | 39.8 | 46.9 | -7.1 | 66.8 | 84.6 | -17.8 | High burden (T3) |
| 292 | 12 | 0.167 | 0.17 | 0.165 | 0.002 | 76.6 | 70.4 | 6.2 | 140.1 | 141.2 | -1.2 | 35 | 40.3 | -5.3 | 49.1 | 56.9 | -7.8 | Low burden (T1) |
| 293 | 13 | 0.061 | 0 | 0.028 | 0.032 | 54.2 | 31.8 | 22.4 | 177.8 | 212 | -34.2 | 32.5 | 33.6 | -1.1 | 57.9 | 71.1 | -13.3 | Low burden (T1) |
| 40 | 16 | 0.149 | 0 | 0.052 | 0.097 | 67.3 | 56.7 | 10.6 | 159 | 173.4 | -14.4 | 33.4 | 33.6 | -0.2 | 53.1 | 58 | -4.9 | Low burden (T1) |
| 41 | 13 | 0.398 | 0.28 | 0.025 | 0.372 | 53.1 | 43.4 | 9.7 | 182.8 | 199.8 | -17 | 35.4 | 35.4 | -0.1 | 64.3 | 70.6 | -6.3 | Mid burden (T2) |
| 42 | 14 | 0.252 | 0.16 | 0.184 | 0.069 | 65.9 | 59.8 | 6.1 | 156 | 153.3 | 2.8 | 36.1 | 41.8 | -5.7 | 56.2 | 63.9 | -7.7 | Mid burden (T2) |
| 43 | 13 | 0.195 | 0.15 | 0.168 | 0.028 | 69.8 | 59.1 | 10.7 | 147.6 | 165 | -17.4 | 39.1 | 38.7 | 0.3 | 57.6 | 63.9 | -6.3 | Low burden (T1) |
| 5 | 10 | 0.13 | 0.1 | 0.064 | 0.067 | 72.5 | 53 | 19.5 | 149.6 | 173.3 | -23.7 | 34 | 34 | 0 | 51 | 58.9 | -7.8 | Low burden (T1) |
| 50 | 13 | 0.492 | 0.39 | 0.432 | 0.061 | 67 | 42.3 | 24.8 | 150.2 | 181.8 | -31.6 | 38.7 | 38.2 | 0.5 | 58 | 68.5 | -10.4 | Mid burden (T2) |
| 55 | 9 | 0.253 | 0.18 | 0.253 | 0 | 82.3 | 67.8 | 14.5 | 132.9 | 152.3 | -19.4 | 32 | 32.2 | -0.2 | 42.6 | 49 | -6.4 | Mid burden (T2) |
| 61 | 17 | 0.36 | 0.31 | 0.129 | 0.231 | 60.2 | 59.2 | 1 | 163.2 | 166 | -2.8 | 40.7 | 44.6 | -3.8 | 66.2 | 73.9 | -7.7 | Mid burden (T2) |
| 67 | 14 | 0.501 | 0.265 | 0.048 | 0.453 | 49.3 | 42.9 | 6.5 | 186.8 | 196.2 | -9.4 | 35.4 | 37.2 | -1.7 | 66.1 | 73 | -6.8 | Mid burden (T2) |
| 77 | 18 | 0.221 | 0.05 | 0.027 | 0.193 | 62.4 | 50.5 | 11.8 | 165.9 | 182.8 | -16.8 | 33.3 | 34.8 | -1.4 | 55.1 | 63.1 | -8 | Mid burden (T2) |
| 78 | 14 | 0.41 | 0.355 | 0.329 | 0.082 | 62.3 | 51.5 | 10.8 | 162.5 | 179.8 | -17.4 | 35.9 | 35.8 | 0.1 | 58.3 | 64.5 | -6.2 | Mid burden (T2) |
| 8 | 13 | 0.215 | 0.19 | 0.114 | 0.102 | 63.8 | 54.1 | 9.7 | 160.7 | 171 | -10.2 | 39.6 | 40.6 | -0.9 | 63.7 | 69 | -5.4 | Mid burden (T2) |
| 82 | 14 | 0.744 | 0.64 | 0.355 | 0.386 | 49.3 | 47.3 | 2.1 | 180.9 | 181.6 | -0.7 | 39.9 | 39.3 | 0.6 | 72.1 | 71.1 | 1 | High burden (T3) |
| 9 | 18 | 0.043 | 0 | 0.043 | 0 | 70.9 | 67.6 | 3.3 | 154.4 | 158.5 | -4.1 | 28.9 | 31.3 | -2.4 | 44.2 | 49.5 | -5.3 | Low burden (T1) |
| 98 | 15 | 0.071 | 0 | 0.016 | 0.055 | 55.5 | 46.4 | 9.1 | 174.6 | 185.5 | -10.9 | 29.1 | 30.3 | -1.2 | 50.7 | 56 | -5.3 | Low burden (T1) |

**Supplementary Table S11.** Baseline HbA1c distributions across the four continuous glucose monitoring datasets included in the upper-limit capping analysis.

| **Dataset** | **Sensor** | **Participants (n)** | **Mean** | **SD** | **Median** |
| --- | --- | --- | --- | --- | --- |
| Aleppo, 2017 | G4 Platinum | 226 | 7.02 | 0.61 | 7 |
| Brown, 2019 | G6 | 168 | 7.5 | 0.93 | 7.5 |
| Lynch, 2022 | G6 | 450 | 7.45 | 0.87 | 7.4 |
| Wadwa, 2023 | G6 | 101 | 7.61 | 1.26 | 7.5 |

**Supplementary Table S12.** Overall Spearman correlations between baseline HbA1c and upper-limit continuous glucose monitoring capping metrics across all participants.

| **Metric** | **Participants (n)** | **Spearman ρ** | **95% CI** | **p value** |
| --- | --- | --- | --- | --- |
| Upper-cap readings (%) | 945 | 0.513 | 0.465-0.559 | <0.001 |
| Total upper-cap time (mins) | 945 | 0.47 | 0.419-0.518 | <0.001 |
| Longest upper-cap run (mins) | 945 | 0.375 | 0.319-0.429 | <0.001 |

**Supplementary Table S13.** Dataset-specific Spearman correlations between baseline HbA1c and upper-limit continuous glucose monitoring capping metrics stratified by study cohort and sensor generation

| **Metric** | **Dataset** | **Sensor** | **Participants (n)** | **Spearman ρ** | **95% CI** | **p value** |
| --- | --- | --- | --- | --- | --- | --- |
| Upper-cap readings (%) | Aleppo_2017 | G4 | 226 | 0.392 | 0.276-0.497 | <0.001 |
|  | Brown_2019 | G6 | 168 | 0.624 | 0.521-0.708 | <0.001 |
|  | Lynch_2022 | G6 | 450 | 0.495 | 0.422-0.562 | <0.001 |
|  | Wadwa_2023 | G6 | 101 | 0.62 | 0.483-0.728 | <0.001 |
| Total upper-cap time (mins) | Aleppo_2017 | G4 | 226 | 0.389 | 0.272-0.494 | <0.001 |
|  | Brown_2019 | G6 | 168 | 0.624 | 0.522-0.708 | <0.001 |
|  | Lynch_2022 | G6 | 450 | 0.458 | 0.382-0.528 | <0.001 |
|  | Wadwa_2023 | G6 | 101 | 0.61 | 0.471-0.72 | <0.001 |
| Longest upper-cap run (mins) | Aleppo_2017 | G4 | 226 | 0.282 | 0.157-0.398 | <0.001 |
|  | Brown_2019 | G6 | 168 | 0.59 | 0.482-0.681 | <0.001 |
|  | Lynch_2022 | G6 | 450 | 0.415 | 0.335-0.489 | <0.001 |
|  | Wadwa_2023 | G6 | 101 | 0.601 | 0.46-0.713 | <0.001 |

**Supplementary table S14.** Comparison of between-device sensor capping metrics derived from Dexcom G4 Platinum (Aleppo) and pooled Dexcom G6 datasets (Brown, Lynch, Wadwa).

| **Metric** | **G4 (n=226) median (IQR)** | **G6 (n=722) median (IQR)** | **Mann–Whitney U** | **p value** | **r (rank-biserial)** |
| --- | --- | --- | --- | --- | --- |
| Upper-cap readings (%) | 0.116 (0.033–0.344) | 0.151 (0.022–0.697) | 75,522 | 0.0909 | 0.074 |
| Lower-cap readings (%) | 0.148 (0.053–0.305) | 0.095 (0.027–0.304) | 90,961 | 0.0091 | -0.115 |
| Total upper-cap time (mins) | 387.5 (110–1126.3) | 300 (50–1197.5) | 86,493.50 | 0.1712 | -0.06 |
| Total lower-cap time (mins) | 457.5 (163.8–1022.5) | 185 (55–553.8) | 104,426 | <0.001 | -0.28 |
| Longest upper-cap run (mins) | 150 (62.5–243.8) | 95 (15–200) | 95,989 | <0.001 | -0.177 |
| Longest lower-cap run (mins) | 75 (45–130) | 40 (15–105) | 102,492 | <0.001 | -0.256 |
| Number of upper-cap runs | 7 (2–18.8) | 8 (2–32) | 77,016 | 0.2023 | 0.056 |
| Number of lower-cap runs | 23.5 (9.3–44.5) | 11 (4–30) | 101,665 | <0.001 | -0.246 |

**Supplementary table S15.** Consistency of time above hyperglycaemia threshold across Dexcom G6 studies.

| **Study** | **n** | **Median (IQR)** |
| --- | --- | --- |
| Brown_2019 | 168 | 0.069 (0.000–0.404) |
| Lynch_2022 | 451 | 0.174 (0.036–0.810) |
| Wadwa_2023 | 103 | 0.177 (0.021–0.743) |

**Supplementary table S16.** Dexcom G6 (Brown, Lynch, Wadwa) comparison Kruskal–Wallis test.

| **Test** | **Metric** | **Statistic** | **df** | **p value** |
| --- | --- | --- | --- | --- |
| Kruskal–Wallis | % upper cap readings | 19.247 | 2 | <0.001 |
