## Supplementary figures for "Prevalence, duration, and clinical implications of Continuous Glucose Monitor (CGM) measurement limit capping in type 1 diabetes"

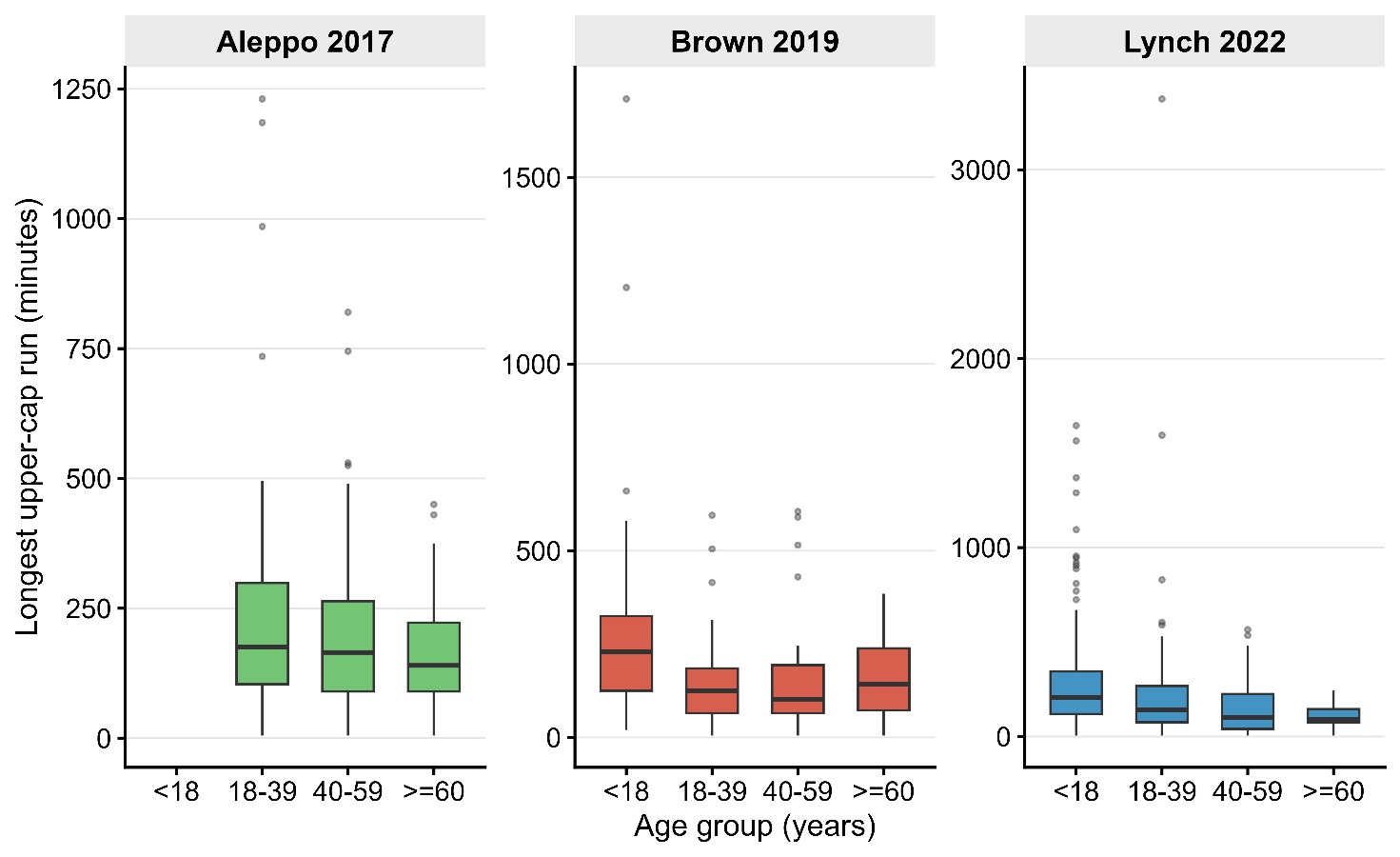

**Supplementary Figure S1. Duration of the longest upper-limit capping run by age group.** Boxplots showing the duration (minutes) of each participant's single longest uninterrupted upper-limit capping run (≥400 mg/dL) by age group (<18, 18–39, 40–59, ≥60 years) in three datasets. Wadwa 2023 is excluded as it enrolled exclusively participants under 18 years. Each box represents the distribution across participants within that age group and dataset; boxes show median and interquartile range, whiskers extend to 1.5×IQR. Y-axis scales are free across datasets.

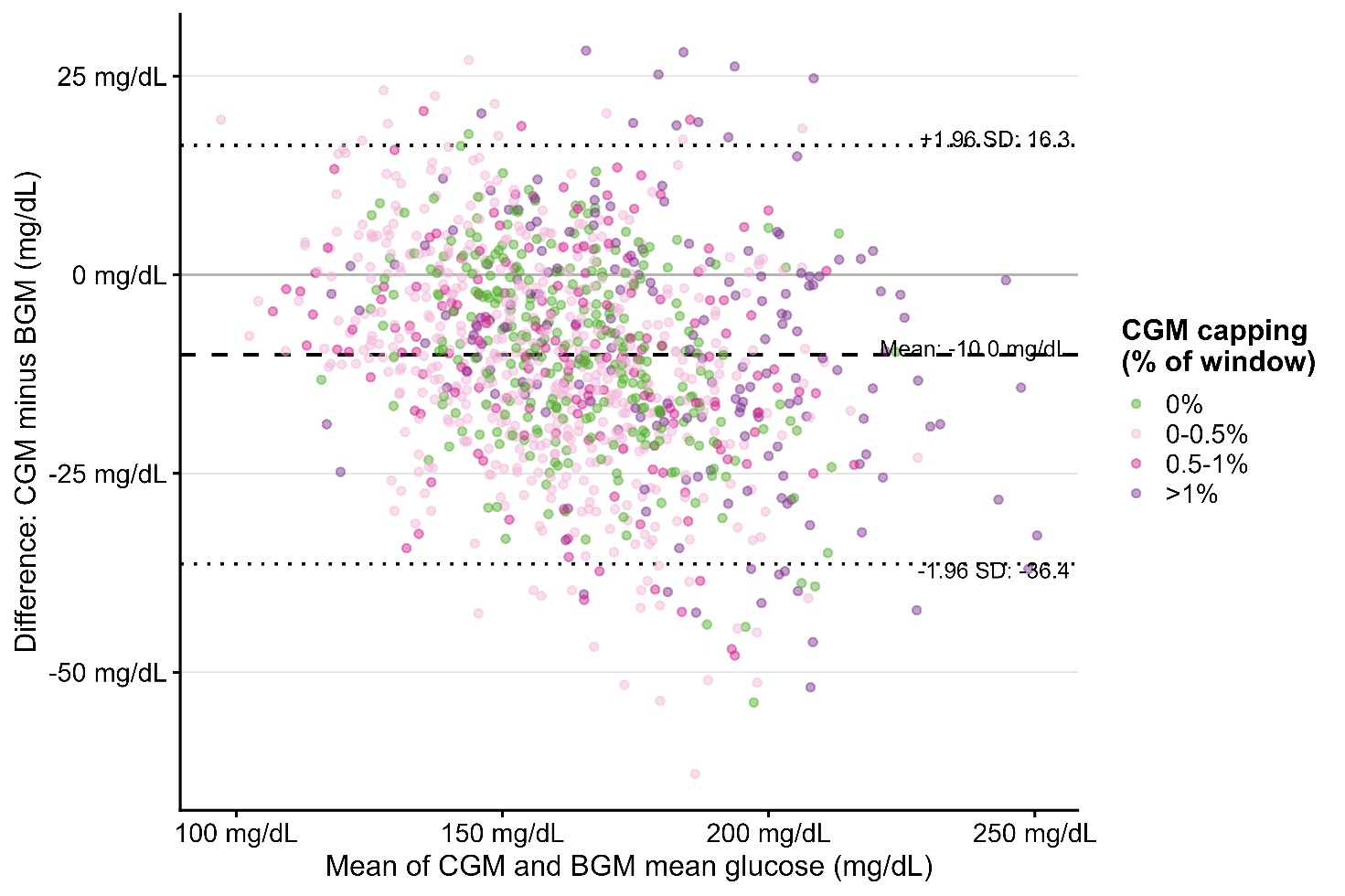

**Supplementary Figure S2. Bland–Altman analysis of agreement between CGM-derived and BGM-derived mean glucose across 14-day windows.** Each point represents one 14-day monitoring window (n=1,162 windows from 77 participants). The x-axis shows the mean of CGM and BGM mean glucose for that window; the y-axis shows the difference (CGM minus BGM). Points are coloured by the proportion of CGM readings within the window that reached a sensor measurement limit (0%, 0–0.5%, 0.5–1%, >1%). Dashed horizontal line indicates the mean bias (CGM − BGM mean difference: −10.0 mg/dL); dotted lines indicate 95% limits of agreement (−36.4 to +16.3 mg/dL). CGM metrics include all readings with capped values retained at their recorded values.

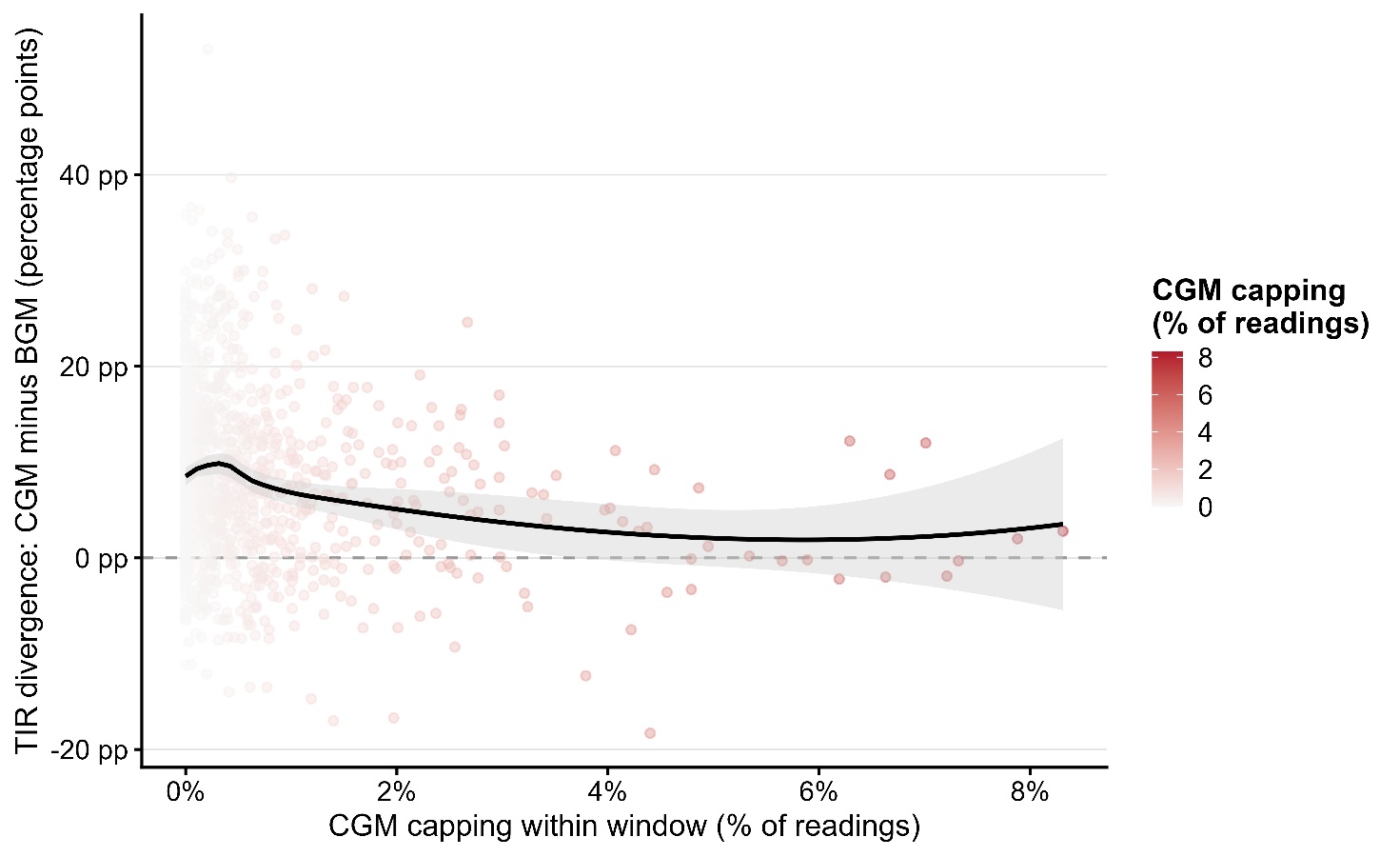

**Supplementary Figure S3. Association between window-level CGM capping burden and time-in-range divergence between CGM and BGM.** Each point represents one 14-day monitoring window (n=1,162 windows from 77 participants). The x-axis shows the percentage of CGM readings within the window that reached a sensor measurement limit (either ≤40 or ≥400 mg/dL); the y-axis shows the difference in time-in-range 70–180 mg/dL (TIR) between CGM and BGM (positive values indicate CGM TIR exceeds BGM TIR). Points are coloured by capping burden. The solid line and shaded region show a LOESS smoother with 95% confidence interval. The absence of a dose-response trend supports the interpretation that CGM capping does not systematically drive TIR overestimation at the prevalence levels observed.

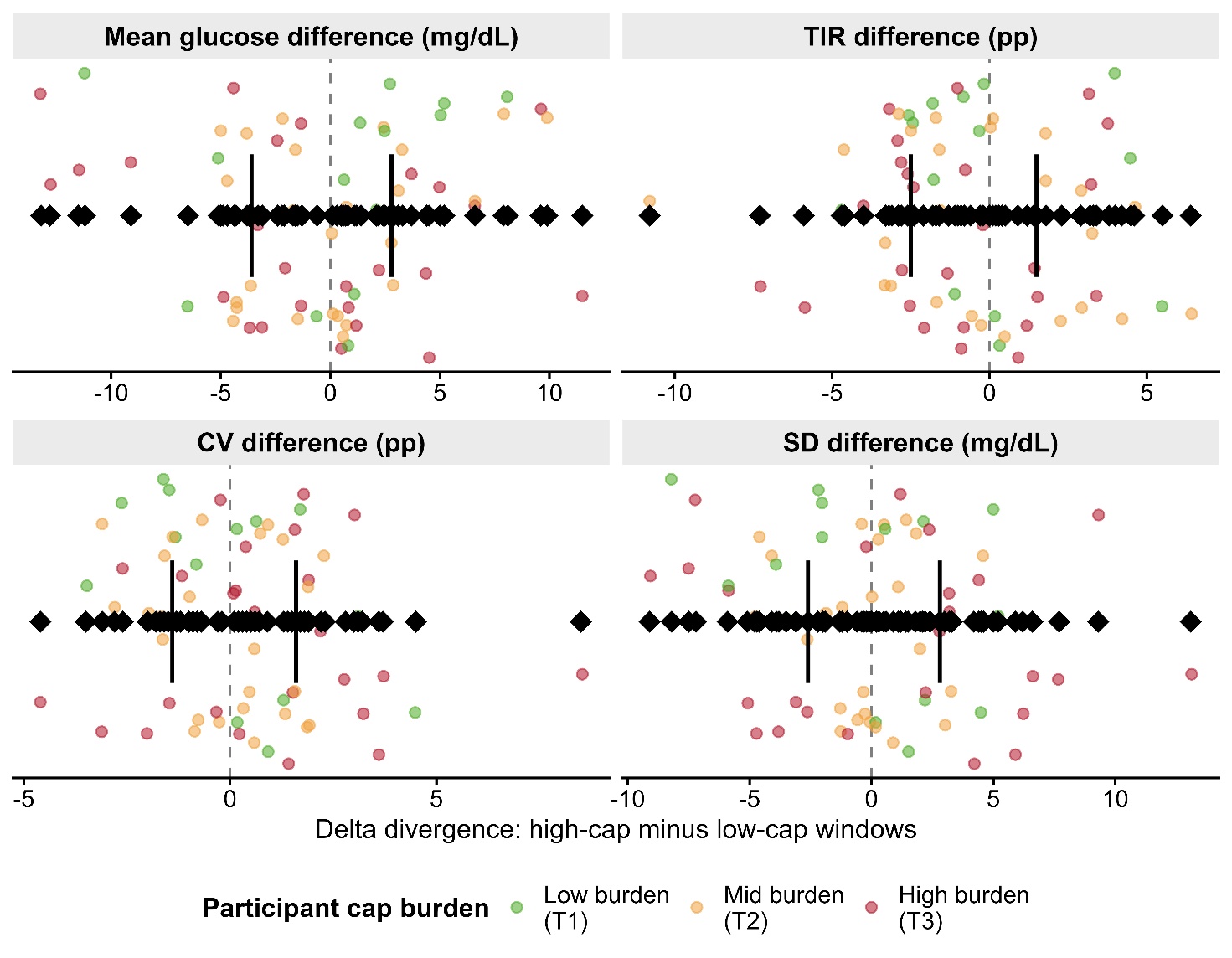

**Supplementary Figure S4. Within-participant effect of CGM capping burden on glucose metric divergence.** Each point represents one participant's difference in CGM–BGM metric divergence between their higher-capping and lower-capping 14-day windows, in the 65 participants with sufficient windows in both categories. For each participant, windows were classified as above or below that participant's own median capping rate; the plotted value is the mean divergence in high-cap windows minus the mean divergence in low-cap windows. Positive values indicate greater CGM overestimation relative to BGM in higher-capping windows. Points are coloured by the participant's overall cap burden tertile (low, mid, high). The diamond symbol and horizontal bar show the overall median and interquartile range. No significant within-participant effect of capping burden was observed for mean glucose (median Δ +0.6 mg/dL, p=0.90), SD (p=0.80), CV (p=0.37), or TIR (median Δ −0.8 pp, p=0.22). A significant effect was observed for TBR <54 mg/dL (p<0.001), although the mechanism underlying this association requires further investigation. pp, percentage points.

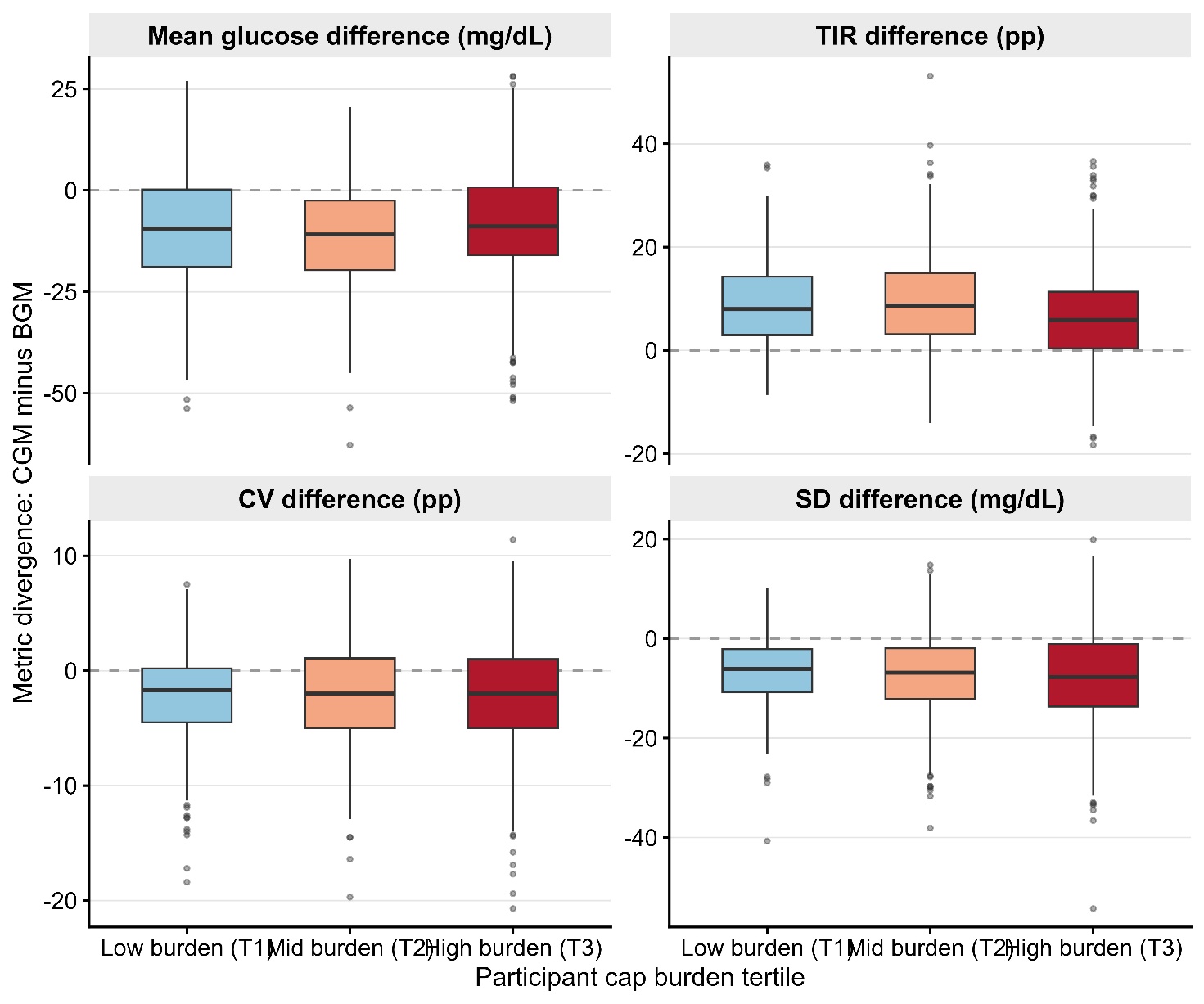

**Supplementary Figure S5. Glucose metric divergence by between-participant CGM cap burden tertile.** Distribution of CGM–BGM metric divergence (CGM minus BGM) across 1,162 14-day monitoring windows, stratified by between-participant CGM cap burden tertile. Participants were classified into tertiles based on their mean capping percentage across all eligible windows: low burden (T1, mean 0.13%, n=26 participants), mid burden (T2, mean 0.38%, n=26), and high burden (T3, mean 1.22%, n=25). Positive values indicate CGM overestimates the metric relative to BGM. Kruskal-Wallis tests showed significant differences across tertiles for TIR (p<0.001) and mean glucose (p=0.007), but not for CV (p=0.96) or SD (p=0.46). The absence of a monotonic dose-response, combined with the null within-participant finding (Figure S4), suggests that between-tertile differences reflect underlying clinical differences between participant subgroups rather than a direct causal effect of capping. Boxes show median and interquartile range; whiskers extend to 1.5×IQR. pp, percentage points.

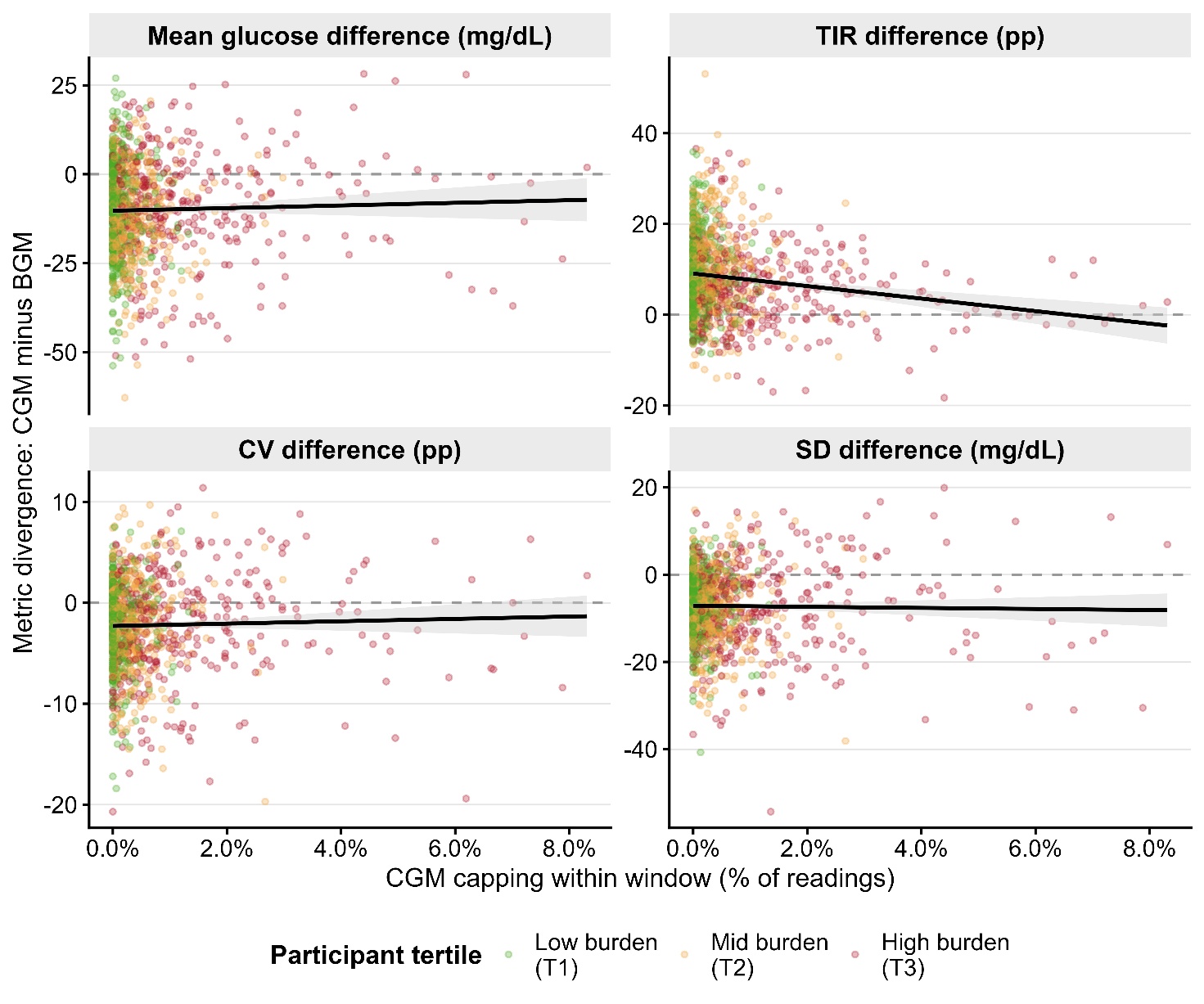

**Supplementary Figure S6. Window-level CGM capping burden and glucose metric divergence by participant tertile.** Association between CGM capping burden (percentage of readings reaching a sensor limit within a 14-day window) and CGM–BGM metric divergence across 1,162 windows from 77 participants. Panels show mean glucose (mg/dL), standard deviation (SD), coefficient of variation (CV), and time in range 70–180 mg/dL (TIR). Points are coloured by the participant's overall cap burden tertile (low T1, mid T2, high T3). Solid lines show linear regression fits with 95% confidence intervals. The overall scatter and flat regression slopes for mean glucose, SD, and CV are consistent with the absence of a systematic dose-response relationship between window-level capping and metric distortion. pp, percentage points.

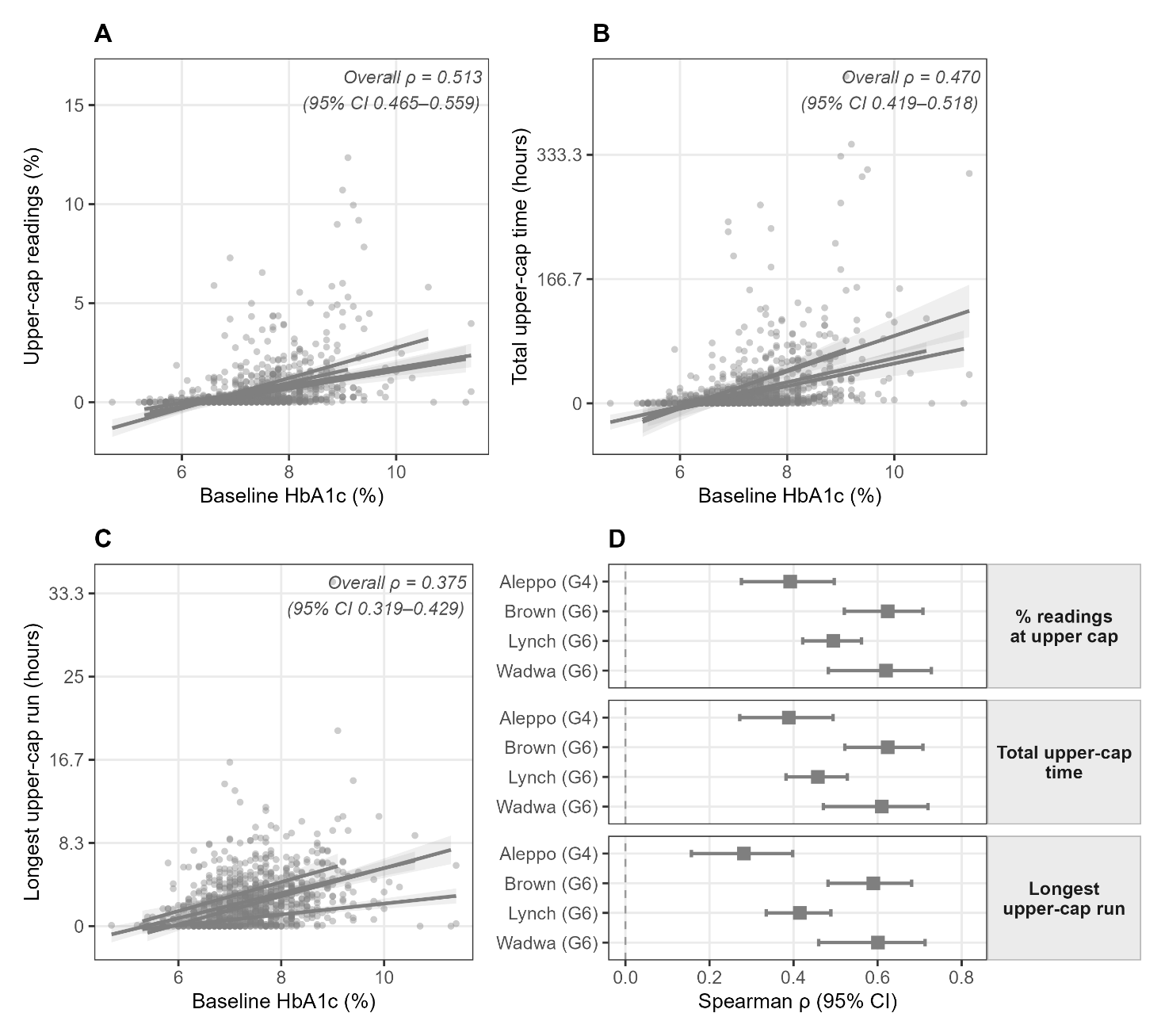

**Supplementary figure S7.** Relationship between baseline HbA1c and upper-limit CGM capping burden across four Dexcom datasets. (A) Percentage of CGM readings recorded at the sensor upper reporting limit, (B) cumulative duration of upper-limit capping, and (C) longest continuous upper-limit capping episode plotted against baseline HbA1c for all participants combined (n=945). Grey points represent individual participants; fitted lines are shown for visualisation only. Overall associations were assessed using Spearman rank correlation.

(D) Dataset-specific Spearman correlation coefficients (ρ) with 95% confidence intervals for each upper-limit capping metric. Aleppo used the Dexcom G4 Platinum sensor, whereas Brown, Lynch, and Wadwa used Dexcom G6 sensors. Positive associations between baseline HbA1c and upper-limit capping burden were observed consistently across all cohorts, indicating that the relationship is not attributable to a specific sensor generation.
