## Supplementary file 2 for "Prevalence, duration, and clinical implications of Continuous Glucose Monitor (CGM) measurement limit capping in type 1 diabetes"

**Supplementary file 2 - Abbreviations**

| Abbreviation | Definition |
| --- | --- |
| AGP | Ambulatory Glucose Profile |
| BGM | Blood Glucose Monitor |
| CGM | Continuous Glucose Monitor |
| CV | Coefficient of Variation |
| Dexcom G4 | Dexcom G4 Platinum Continuous Glucose Monitoring System |
| Dexcom G6 | Dexcom G6 Continuous Glucose Monitoring System |
| GMI | Glucose Management Indicator |
| HbA1c | Glycated Haemoglobin |
| IQR | Interquartile Range |
| MARD | Mean Absolute Relative Difference |
| mg/dL | Milligrams per Decilitre |
| mmol/L | Millimoles per Litre |
| mmol/mol | Millimoles per Mole |
| N | Number/sample size |
| NICE | National Institute for Health and Care Excellence |
| SD | Standard Deviation |
| TAR | Time Above Range |
| TBR | Time Below Range |
| TIR | Time In Range |
| ≥ | Greater than or equal to |
| ≤ | Less than or equal to |
| ± | Plus or minus |
