## Supplementary file 1 for "Prevalence, duration, and clinical implications of Continuous Glucose Monitor (CGM) measurement limit capping in type 1 diabetes"

**Wadwa 2023 Dexcom Clarity export structure**

**Background**

The Wadwa 2023 (PEDAP) dataset showed a markedly different pattern of upper-limit capping runs compared with the other three datasets: 28,403 upper-cap runs with a median duration of only 5 minutes and just 1.2% lasting ≥15 minutes, compared with medians of 35–40 minutes and >70% lasting ≥15 minutes in Brown, Lynch, and Aleppo. This divergence is not consistent with a genuine biological difference in glucose behaviour and requires explanation.

**Investigation**

Inspection of raw Wadwa CGM data revealed that the majority of consecutive upper-cap readings (≥400 mg/dL) were separated by single-reading gaps, where one non-capped reading appeared between two capped readings, thereby breaking what would otherwise be a single sustained capping run into multiple short runs. This pattern was distributed across participants and across the full duration of the study, rather than clustering around particular participants or time periods.

We propose that the most plausible explanation for this is that the Wadwa dataset was exported from Dexcom Clarity in a manner that periodically re-uploaded CGM data in shorter segments. In this scenario, each re-upload segment begins with the device's last transmitted reading, which may fall at or near the capping boundary, before continuing with readings from the new segment. This creates apparent single-reading "interruptions" at the boundaries between segments that do not correspond to true physiological transitions out of the capped range, with the result that it fragments real sustained capping runs into many short 5 minute runs.

This interpretation is supported by our deduplication step for the Wadwa dataset, which removed 843,810 timestamp conflict records, a substantially higher number than forthe other datasets (955 exact duplicates across all datasets). The large number of timestamp conflicts is consistent with repeated re-uploading of overlapping data segments from Clarity.

**Implications for analysis**

Because the Wadwa cap run structure reflects data export artefact rather than genuine glucose physiology, Wadwa run duration data were excluded from cross-dataset summaries of capping run duration (Table 2 and Figure 2). Capping prevalence data (proportion of readings at each limit, proportion of participants affected) are not affected by this export structure and are reported normally for Wadwa across all analyses. Age and sex association analyses also use participant-level capping prevalence metrics rather than run durations and so are also unaffected.

Users of the PEDAP public dataset who wish to analyse capping run durations should be aware of this export characteristic and consider pre-processing steps to merge runs separated by single non-capped readings before performing run duration analyses.
